# A Comprehensive Review of Iron Prophylaxis in the National Anemia Control Programme in India (Anemia-Mukt Bharat)

**DOI:** 10.64898/2026.01.29.26345166

**Authors:** Subhanwita Manna, Ranadip Chowdhury, Raghu Pullakhandam, Tanica Lyngdoh, K. Madhavan Nair, Vani Kandpal, Kapil Yadav, Molly Jacob, Abhishek Jaiswal, Priyanka Gupta Bansal, Prashanth Thankachan, Reema Mukherjee, Bharati Kulkarni

## Abstract

Anaemia remains a significant public health issue in India, despite five of control programs. Anaemia affects 52-67% of target populations in India despite five decades of control programmes. We conducted a review of reviews and meta-analyses (SRMAs) of regional studies to evaluate daily versus intermittent oral iron and iron-folic acid (IFA) supplementation across age groups. We identified 21 SRMAs (17 high-quality, 4 moderate-quality) and 44 regional studies from India and South Asia. IFA prophylaxis consistently improved haemoglobin levels (4.1-8.8 g/L increase) and ferritin concentrations, reducing anaemia risk by 23-70% across all age groups. IFA prophylaxis consistently improved haemoglobin levels (4.1-8.8 g/L increase) and ferritin concentrations, reducing anaemia risk by 23-70% across all age groups. Daily and intermittent regimens showed similar haematological outcomes in children, adolescents, and women of reproductive age. Among pregnant women, daily supplementation was superior for haemoglobin and ferritin levels, though intermittent dosing had fewer gastrointestinal side effects. These findings support weekly IFA supplementation for non-pregnant beneficiaries as an evidence-based strategy, even in settings where anaemia prevalence exceeds 40%. Further research on long-term safety in non-iron-deficient populations is needed.

## INTRODUCTION

Anemia affects an estimated 57% of women and girls of reproductive age (WRA) (15-49 years), 67% of children under 5 years of age, and 52% of pregnant women in India (1). Iron deficiency is considered to be the major cause in LMICs, accounting for about 50% of anemia in school children and WRA, and 80% in children aged 2–5 years (2,3) Anemia may lead to impaired cognitive development, reduced productivity, and increased adverse outcomes for the mother and fetus(4). Therefore, effective public health strategies to address anaemia have been a national priority.

Anemia control programs have been in place in India for several decades (5–7). However, a comparison of NFHS-4 (7) and NFHS-5(1) data shows little to no improvement across all age groups: pregnant women (50.4% to 52.2%), children (58.6% to 67.1%), men (22.7% to 25%), women (53.1% to 57%), and adolescents (boys: 29.2% to 31.1%; girls: 54.1% to 59.1%)(8). This stagnation despite substantial programmatic investment suggests that current strategies may require re-evaluation.

The Indian Council of Medical Research (ICMR) evaluated specific aspects of the programme, particularly targeted beneficiaries, diagnostic methods, and prophylaxis and treatment strategies. The currently available evidence supporting AMB programme interventions were reviewed and areas where modifications or newer interventions might enhance effectiveness were identified. Scientists, clinicians, academicians, programme experts and officials from involved ministries, reviewed the existing evidence, focussing on relevant research questions.

## PURPOSE OF THE CURRENT REVIEW

Oral iron-folic acid (IFA) prophylaxis is a cornerstone of the AMB program, addressing the major causes of anemia (iron and folate deficiency). The AMB guidelines(9) differ from the WHO recommendations (10–13) regarding doses and regimens used for oral iron prophylaxis (Supplementary Table 1). In summary, WHO recommends daily iron prophylaxis for three months in a year for children, menstruating women, and pregnant women when the prevalence of anaemia is 40% or higher. In regions where the prevalence is between 20–40%, intermittent prophylaxis is recommended for three months followed by a three-month break after which the regimen is repeated. The ICMR and National Institution for Transforming India (NITI) Aayog convened meetings involving public health specialists, government officials, scientists, clinicians, academicians and experts in the field, nutritionists, haematologists, and AMB program stakeholders. A comprehensive review was conducted to identify potential areas to focus on for improvement in strategies for prophylaxis, treatment protocols, target population, and program design Given that universal supplementation programmes reach both iron-deficient and non-deficient individuals, understanding the safety profile in the latter group is crucial. A working group reviewed evidence related to IFA prophylaxis. Through iterative feedback, group discussions, and expert validation, consensus was reached on final recommendations.

This article summarises the evidence review undertaken by the working group, addressing three key questions:

Research Question 1: How effective is iron/iron-folic acid (IFA) supplementation in prophylactic doses, compared to no supplementation, in reducing the prevalence of anaemia, iron deficiency, and iron-deficiency anaemia, as well as in improving non-hematological outcomes in children (aged 6 months to 59 months, and 5 to 9 years), adolescents (aged 10 to 19 years), women of reproductive age (aged 15 to 49 years), and pregnant women?

Research Question 2: Is daily oral iron supplementation in prophylactic doses more effective than intermittent supplementation in reducing the prevalence of anemia, iron-deficiency anemia, and iron deficiency among children (aged 6–59 months, and 5–9 years), adolescents (aged 10–19 years), women of reproductive age (aged 15–49 years), and pregnant women?

Research Question 3: What are the potential adverse effects of oral IFA supplementation given in prophylactic doses, especially in non-iron deficient individuals?

## METHODOLOGY

### Overview

This review comprised two complementary components: (i) a systematic review of existing SRMAs to synthesise global evidence, and (ii) a meta-analysis of studies from India and South Asia to generate region-specific estimates.

### Part 1: Review of Systematic Reviews and Meta-analysis

#### Search Strategy

We searched PubMed and the Cochrane Library from inception to December 2024 for systematic reviews and meta-analyses examining iron prophylaxis effects in AMB target groups. Search terms combined concepts of population (infants, children, adolescents, women of reproductive age, pregnant women), intervention (iron supplementation, daily, intermittent), and outcomes (anaemia, adverse effects). No language restrictions were applied. Reference lists of relevant articles were hand-searched for additional studies. Detailed search strategies are provided in Supplementary Table 2.

#### Eligibility Criteria

We included: Systematic reviews and meta-analyses of randomised controlled trials (RCTs) or quasi-randomised trials - Studies examining oral iron supplementation using various iron salts at prophylactic doses - Reviews addressing: (i) haematological and non-haematological effects of iron/IFA supplementation, (ii) comparisons of intermittent versus daily dosing, and (iii) adverse effects across age groups, including individuals without anaemia or iron deficiency. For Question 3 (adverse effects), due to limited evidence, we also included RCTs and observational studies from India and SRMAs incorporating observational studies. The study protocol is registered in PROSPERO (CRD42025642650).

#### Study Selection

Two reviewers independently screened titles and abstracts using Rayyan software (13), followed by full-text review for inclusion. Disagreements were resolved through discussion or involvement of a third reviewer.

#### Quality Assessment

Methodological quality of reviews was assessed using the AMSTAR tool(15), categorizing them as high (scored: 8–11), moderate (Scored: 4–7), or low (Scored: 0–3) quality. Two reviewers (SM and VK) independently evaluated the reviews, with disagreements resolved by a third author (RM). AMSTAR ratings are presented in Supplementary Table 3

#### Data extraction

Data were extracted into a standardised template including: study characteristics (authors, year, target population, sample size), intervention details (dose, duration, frequency), comparison groups, outcomes (haemoglobin, ferritin, anaemia prevalence, iron deficiency, IDA, adverse effects), and quality ratings.

### Part 2: Meta-analysis of regional studies

#### Study Identification

From shortlisted SRMAs, we identified studies conducted in India and South Asia. An additional search was conducted to identify studies not included in the shortlisted SRMAs, using the same keyword combinations. We included only studies that specifically addressed any of the three research questions. Studies were categorised by target groups: (i) preschool children (6-59 months), (ii) children (5-9 years), (iii) adolescents (10-19 years), (iv) non-pregnant WRA (≥15 years), and (v) pregnant women.

#### Risk of Bias assessment

Quality of individual trials was assessed using criteria from the Cochrane Handbook for Systematic Reviews of Interventions. Each domain was rated as low risk, unclear risk, or high risk of bias. For quantitative analysis, we assigned codes 0, 1, and 2 for low, unclear, and high risk respectively. An overall quality score was generated by summing domain-specific scores. Two reviewers independently assessed risk of bias, resolving disagreements through discussion with a third reviewer.

#### Data Extraction

Two reviewers independently extracted data using a form developed for this review, including: study characteristics (author, year, country, sample size, participant characteristics), intervention details (dose, duration, frequency), and outcomes (mean ± SD for haemoglobin (g/L) and ferritin (µg/L) pre- and post-intervention). Where means and standard deviations were not reported directly, we derived them using established statistical methods based on available median and interquartile range or other summary statistics. The data extraction templates were piloted on four studies and refined prior to full extraction. Discrepancies were resolved through discussion at each stage. Titles and abstracts were screened independently using Rayyan software(15) by two reviewers, followed by a full-text review for inclusion. Data were extracted into a standardized template and analyzed using STATA v18. Effect sizes were estimated using a random-effects model, and forest plots were generated.

#### Statistical Analysis

A meta-analysis assessed efficacy of iron interventions on haemoglobin and ferritin levels. Mean differences (MDs) with 95% confidence intervals (CIs) were calculated using a random-effects model to account for expected heterogeneity across populations, settings, and interventions. Heterogeneity was assessed using I² statistics, and tau² was estimated using the restricted maximum-likelihood method. Publication bias was evaluated visually through inspection of funnel plots and statistically using Egger’s regression test. Funnel plots were generated when more than 10 studies were available for an outcome, following methodological guidance. Subgroup analyses were performed for each primary outcome (haemoglobin and ferritin levels) based on age group of participants.

Meta-regression was conducted to explore potential sources of heterogeneity for each primary outcome separately. Study-level covariates assessed included sample size, intervention duration, intervention dosage, and quality score. All analyses were performed using STATA version 18.

#### Reporting

Data extraction and reporting adhered to PRISMA guidelines (Supplementary Figure 2).

## RESULTS

### Overview of included studies

We identified 593 systematic reviews from PubMed and Cochrane databases. After screening 58 full-texts, 20 reviews (19 SRMAs, and 1 SR) were included in the final analysis (Figure 1, Supplementary Material). Among these, 15 were rated high quality and 5 moderate quality by AMSTAR assessment(16). Results are detailed in Supplementary Tables 4–6, summarizing authors, target groups, sample sizes, interventions& control, adherence/duration & doses, and outcomes.

**Figure 1:**
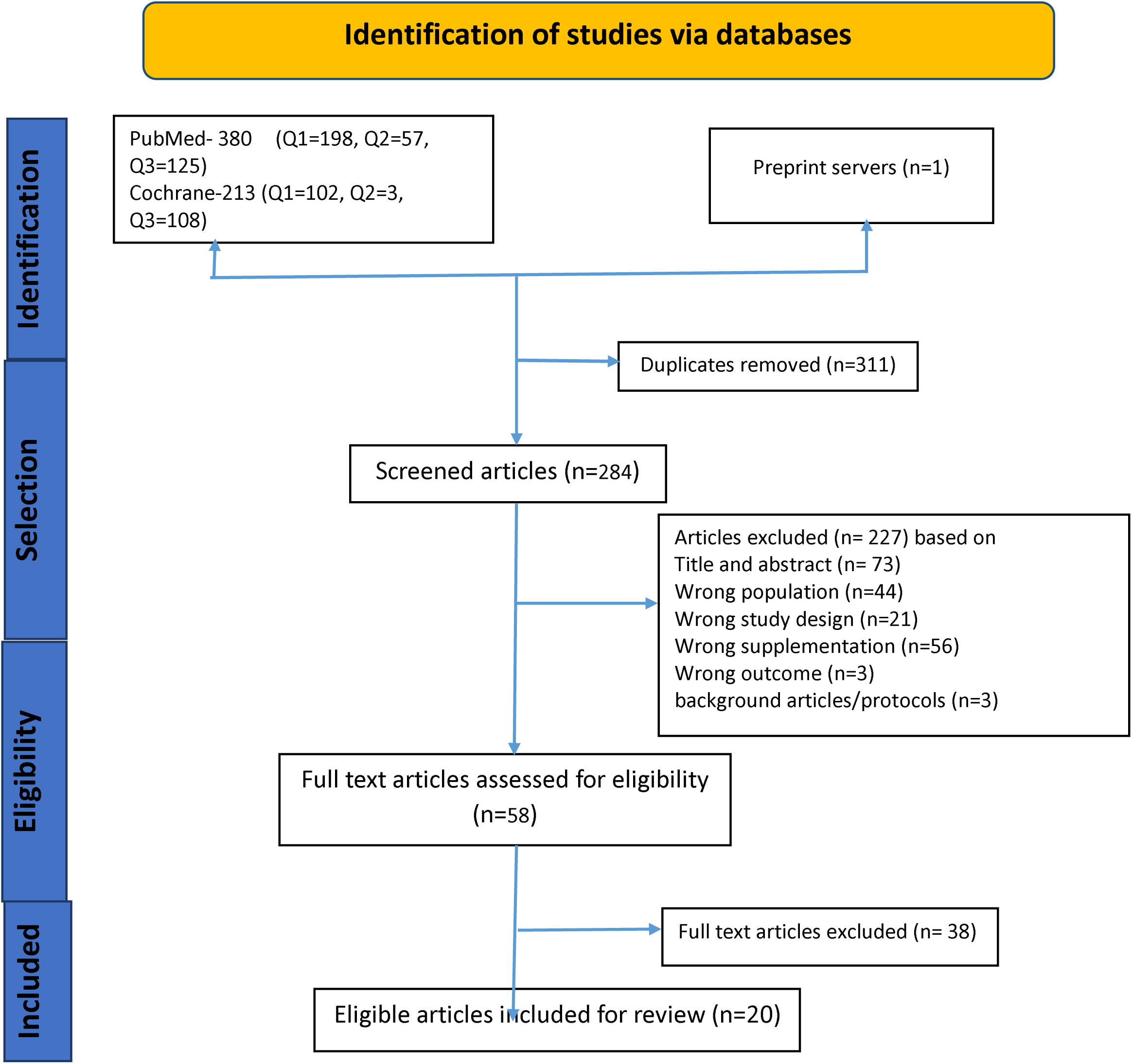
PRISMA flow chart.

### Regional Studies from India and South Asia

#### Study characteristics

From the 20 reviews we identified 41 studies conducted in India and other South Asia. An additional 03 studies were identified through an updated search, yielding a total of 44 studies. We found 24 studies from India (17–39), 06 from Pakistan (40–46) and Bangladesh (47–53) each, 07 studies from Sri Lanka(54–59) and one from Nepal(60). Distribution by age group: 13 studies in each in children 6-59 months and school-age children (5-9 years), 04 studies in adolescents(20,27–29, 36,38,43, 59–64), 01 in WRA(55) and 11 in pregnant women (23,31,33,36,41,42,45,49,51,56). One study specifically reported adverse effects of iron/IFA supplementation in adolescents(38). Details of the studies were reported in table no 1.

### Intervention characteristics

#### Children 6-59 months

Iron supplementation was administered daily or intermittently, with doses ranging from 2 mg/kg to 100 mg and durations from 8 weeks to 18 months. Daily dosing was most used.

#### Children 5-18 years

Both daily and intermittent schedules were employed, with dosages ranging from 7.5 mg/kg to 100 mg over 2-12 months.

#### Adolescents

Supplementation doses varied from 50 mg to 100 mg for 10 weeks to 8 months. Some trials combined intermittent dosing with vitamin A or educational programmes.

#### WRA

Only one study reported iron prophylaxis with a daily dose of 200 mg for 2 months.

#### Risk of bias assessment

The methodological quality across seven domains is presented in Figure 8-9. Overall, the majority of studies had a low to moderate risk of bias, with performance bias and allocation concealment contributing most to the potential methodological limitations.

##### RESEARCH QUESTION 1- Efficacy of Iron/IFA supplementation

Seventeen reviews examined haematological and other outcomes of IFA prophylaxis : 10 on children and adolescents(65–74) , 02 on WRA(75,76) and 06 on pregnant women (one also included lactating women and children)(72,77–81). Interventions involved daily or intermittent iron prophylaxis, compared to placebo, no supplement, or non-iron supplements. While studies varied in age group definitions, results are categorized by AMB program beneficiary groups. Results are summarised in Supplementary table 4

### Haemoglobin Levels

#### Children 6-59 months

Iron supplementation significantly increased haemoglobin levels across multiple high-quality reviews: - MD 4.1 g/L (95% CI: 2.8, 5.3 g/L; age 6-23 months)(72) – MD 7.22 g/L (95% CI: 4.87, 9.57 g/L; age 4-23 months)(71) - MD 6.02 g/L (95% CI: 4.28, 7.76 g/L; age <5 years) (24) - WMD 5.7 g/L (95% CI: 3.8, 7.7 g/L; age 2-<5 years)(65)

#### Children 5-9 years and adolescents (10-19years)

Haemoglobin improvements with supplementation were: MD 8.38 g/L (95% CI: 6.21, 10.56 g/L; age 5-12 years)(70), SMD 1.08 (95% CI: 0.68, 1.49; age 6-12 years)(69) - MD 5.20 g/L (95% CI: 2.51, 7.88 g/L; age <12 years)(67) - WMD 7.4 g/L (95% CI: 6.1, 8.7 g/L; age <18 years)(68) - WMD 6.6 g/L (95% CI: 4.4, 8.7 g/L; age >12 years) (65) IFA prophylaxis led to greater haemoglobin improvements in anaemic children compared to non-anaemic children (65,68,71).

#### WRA

Reported improvements were: - MD 5.30 g/L (95% CI: 4.14, 6.45 g/L)(75) - MD 5.19 g/L (95% CI: 3.07, 7.32 g/L)(76)

#### Pregnant women

One review reported MD in haemoglobin at term of 8.83 g/L (95% CI: 6.55, 11.11 g/L)(82)

### Anemia Prevalence

Across multiple high-quality reviews, iron supplementation significantly reduced anaemia risk across all age groups (Supplementary Table 5).

#### Children 6-59 months

- 39% reduction (RR 0.61, 95% CI: 0.50, 0.74; age 4-23 months; high quality)(71) - 45% reduction (RR 0.55, 95% CI: 0.44, 0.70; age <5 years; high quality)(74) - 23% reduction (RR 0.77, 95% CI: 0.67, 0.90; age 6-23 months; high quality)(72) - 24% reduction (RR 0.76, 95% CI: 0.53, 1.09; age 2-<5 years; high quality)(65). Tam et al. reported greater reduction in non-anaemic under-five children at baseline than in anaemic children(74).

#### Children 5-9 years and adolescents 10-19 years

51% reduction (RR 0.49, 95% CI: 0.31, 0.79; age >12 years; high quality)(65), 50% reduction (RR 0.50, 95% CI: 0.39, 0.64; age 5-12 years; high quality)(70) - 49% reduction (RR 0.51, 95% CI: 0.37, 0.72; age <12 years; high quality)(67)

#### WRA

61% reduction (RR 0.39, 95% CI: 0.25, 0.60)(75), 35% reduction (RR 0.65, 95% CI: 0.49, 0.87; high quality)(76)

#### Pregnant women

70% reduction (RR 0.30, 95% CI: 0.19, 0.46; high quality)(78),70% reduction (RR 0.30, 95% CI: 0.20, 0.47; high quality)(79)

High-quality studies reported anaemia risk reduction of 23-45% in young children (65,74), 49-51% in older children (65,67,70), 35-61% in WRA(75,76), and 70% in pregnant women(79).

### Ferritin Levels

#### Children 6-59 months

- MD 21.42 ng/mL (95% CI: 17.25, 25.58 ng/mL; age 4-23 months; high quality)(71) , MD 17.3 ng/mL (95% CI: 13.5, 21.2 ng/mL; age 6-23 months; moderate quality)(72) , WMD 14.4 ng/mL (95% CI: 8.7, 20.0 ng/mL; age 6-23 months; high quality)(65)

#### Children 5-9 years and adolescents 10-19 years

MD 28.45 ng/mL (95% CI: 18.03, 38.86 ng/mL; age 5-12 years; high quality)(70) , WMD 10.9 ng/mL (95% CI: 9.1, 12.7 ng/mL; age >12 years; high quality)(65) - One review showed no effect (SMD 1.93; 95% CI: -0.27, 4.14; age 6-12 years; moderate quality)(69), Pasricha et al.(71) found greater ferritin increases in anaemic children, while Andersen et al. (65) reported higher gains in mixed or non-anaemic groups.

#### WRA

MD 10.27 ng/mL (95% CI: 8.90, 11.65 ng/mL; high quality) (75) , MD 7.46 ng/mL (95% CI: 5.02, 9.90 ng/mL; high quality)(76)

All age groups showed consistent ferritin improvements with iron supplementation. Most high-quality studies in children and adolescents reported clear benefits, though one moderate-quality study found no effect (69).

### Iron Deficiency (ID) and Iron Deficiency Anaemia (IDA)

Oral iron supplementation markedly reduced risk for both ID and IDA across all age groups.

#### Children <5 years

Iron Deficiency: 62% reduction (RR 0.38, 95% CI: 0.27, 0.53; age 6-23 months; high quality) (65) , 70% reduction (RR 0.30, 95% CI: 0.15, 0.60; age 4-23 months; high quality)(71) , 69% reduction (RR 0.31, 95% CI: 0.14, 0.67; age 2-<5 years; high quality)(65) , 79% reduction (RR 0.21, 95% CI: 0.12, 0.39; age <5 years; high quality)(74) Iron Deficiency Anaemia: 68% reduction (RR 0.32, 95% CI: 0.14, 0.69; age 6-23 months; high quality)(65) , 86% reduction (RR 0.14, 95% CI: 0.10, 0.22; age 4-23 months; high quality) (71) , 86% reduction (RR 0.14, 95% CI: 0.04, 0.54; age <5 years; high quality)(74) , 81% reduction (RR 0.19, 95% CI: 0.06, 0.56; age 2-<5 years; high quality)(65)

### Older children and adolescents

Iron Deficiency: 79% reduction (RR 0.21, 95% CI: 0.07, 0.63; age 5-12 years; high quality)(70) , 70% reduction (RR 0.30, 95% CI: 0.11, 0.80; age >12 years; high quality)(65) , 76% reduction (RR 0.24, 95% CI: 0.06, 0.91; age <12 years; high quality)(67)

Iron Deficiency Anaemia: 91% reduction (RR 0.09, 95% CI: 0.01, 1.64; age >12 years; high quality)(65)

Andersen et al. reported higher ID and IDA risk reduction in those anaemics at baseline (65).

#### WRA

Iron Deficiency: 38% reduction (RR 0.62, 95% CI: 0.50, 0.76; high quality)(75) and 50% reduction (RR 0.50, 95% CI: 0.24, 1.04; high quality)(76)

#### Pregnant women

ID at term: 57% reduction (RR 0.43, 95% CI: 0.27, 0.66; high quality)(78) , IDA at term: 77% reduction (RR 0.33, 95% CI: 0.16, 0.69; high quality)(78)

Across all age groups, oral iron supplementation consistently reduced risk of iron deficiency (60-80%) and iron-deficiency anaemia (70-90%). High-quality studies demonstrated strong and sustained benefits(65,70,71,74), with greater reductions among participants who were anaemic at baseline(65).

### Growth, Cognitive and Fetal Outcomes

#### Growth

##### Children 6-59 months

Three reviews (71,73,74) reported no effect of iron supplementation on growth.

##### Children 5-9 years and adolescents 10-19 years

One review(70) reported no benefit on growth outcomes. Andersen et al. reported increased height-for-age Z-scores in anaemic children (RR 0.20, 95% CI: 0.01, 0.40; age <20 years; high quality) but no effect in non-anaemic children or mixed groups (65).

##### WRA

Daily oral supplement had no effect on weight and BMI(75).

### Cognitive Outcomes

#### Children 4-23 months

Pasricha et al. reported improvement in mental development index (MD 4.46, 95% CI: -9.32, 18.24), with anaemic children benefitting most (71).

#### Children 5-9 years and adolescents 10-19 years

Iron supplementation improved memory (Digit Span test, Visual Memory Test, Hopkins Verbal Learning Test): SMD 0.47 (95% CI: 0.13, 0.81; age 6-12 years; moderate quality)(69), Intelligence scores SMD 1.01 (95% CI: 0.34, 1.68; age 6-19 years; high quality)(66) and SMD 0.79 (95% CI: 0.41, 1.16; age 6-12 years; moderate quality) (69). Benefits were particularly evident among those who were anaemic at baseline.

### Foetal Outcomes

Minimal effects on foetal outcomes such as low birth weight, preterm birth, or neonatal deaths were observed (Supplementary Table 4)(78,79).

High-quality evidence showed iron supplementation had no effect on growth/anthropometry in children and WRA (65,70,71,74,75), with similar findings in moderate-quality studies(73). Improvements in memory and intelligence were consistently reported, particularly among anaemic children, supported by moderate- and high-quality evidence (66,69).No effect on foetal outcomes were reported

### Regional Data from India and South Asia

#### Haemoglobin Concentration

Regional studies showed an overall increase in haemoglobin of MD 6.51 g/L (95% CI: 4.14–8.88 g/L; 29 studies). A subgroup analysis by age group reported the following: children aged 6–59 months, MD 6.31 g/L (95% CI: 2.03–10.60 g/L; 13 studies)(16,17,25,20,26,29,31,32,39,42,47,51); children aged 5–9 years, MD 7.68 g/L (95% CI: 3.55–11.81 g/L; 11 studies) (17–19,22,24,25,34,40,43,48,58)(52)(26); adolescents, MD 8.42 g/L (95% CI: 4.65–12.18 g/L; 4 studies)(35,57,59)(47); and adolescent girls, MD 6.53 g/L (95% CI: 1.57–11.49 g/L; 2 studies)(58,51).

#### Serum Ferritin Levels

Across 17 studies, iron supplementation was associated with a pooled mean increase in ferritin of 12.49 ng/mL (95% CI: 7.10,17.87 ng/mL). Subgroup analysis by age group showed a significant improvement among children aged 5–9 years (MD: 19.60 ng/mL; 95% CI: 10.34,28.86 ng/mL; 3 studies), followed by children 6-59 months- (MD: 12.74 ng/mL; 95% CI: 3.71,21.78 ng/mL; 10 studies), and adolescents (MD: 7.86 ng/mL; 95% CI: 1.48,14.25ng/mL; 4 studies).

#### Meta-Regression Analysis

Meta-regression analyses were performed separately for haemoglobin and ferritin outcomes to identify potential sources of heterogeneity. For haemoglobin, both total sample size (β = 0.004, *p* = 0.01) and intervention duration (β = 0.059, *p* = 0.017) were found to be significant predictors of effect size, with residual heterogeneity decreasing marginally (I² = 99.99% to 98.91%). For ferritin, total sample size (β = 0.031, *p* = 0.02) showed a significant contribution, resulting in a slight reduction in residual heterogeneity (I² = 99.65% to 99.16%). Overall, these covariates accounted for only a small proportion of the between-study variability. The persistently high I² values indicate that additional unmeasured factors—such as baseline iron status, co-existing nutritional deficiencies, inflammation, adherence, population characteristics, and variations in outcome measurement—likely contributed to the observed heterogeneity.

#### Publication Bias

For both haemoglobin and ferritin, funnel plots showed visual heterogeneity. However, Egger’s test showed no statistically significant evidence of publication bias (p = 0.21 for haemoglobin; p = 0.11 for ferritin).

### Summary of evidence for Research Question 1 (Efficacy of iron/ iron folic acid supplementation)

Across 17 systematic reviews (13 high-quality, 4 moderate-quality), prophylactic oral iron supplementation consistently improved haematological outcomes across all population groups. Haemoglobin concentrations increased by 4-7 g/L in young children (65,71,72,74), 5-8 g/L in older children and adolescents (65,67–70), approximately 5 g/L in WRA(75,76), and 8.8 g/L at term in pregnant women(82).

Anaemia risk: High-quality reviews reported reductions of 23-45% in younger children(65,74), 49-51% in older children(65,67), 35-61% in WRA(75,76), and 70% in pregnant women(79).

Ferritin: Consistent improvements across all age groups. High-quality studies demonstrated clear benefits(65,70–72,74,75), though evidence regarding baseline anaemia status was mixed(65,71).

Iron deficiency and IDA: Supplementation consistently reduced ID risk by 60-80% and IDA risk by 70-90% across all age groups, with consistent effects replicated across high-quality reviews(65,70,71,74). Greater reductions were observed among anaemic participants(65). In WRA, ID risk decreased by 38-50%(75,76), and in pregnant women, ID and IDA risks decreased by 57% and 77% respectively (78).

Growth, cognitive and adverse birth outcomes: High-quality evidence showed no measurable effect on growth in children or WRA(65,70,71,74,75), consistent with moderate-quality findings(73).Cognitive outcomes: Demonstrated clearer benefits, with improvements in memory and intelligence consistently observed, particularly among anaemic children, supported by moderate- and high-quality evidence(66,69).Foetal outcomes: Among pregnant women, effects were minimal, with limited impact on low birth weight, preterm birth, and neonatal mortality (78,46).

#### Regional data

In India and South Asia, supplementation improved haemoglobin across all target groups, with adolescents showing greatest gains and ferritin improvements most evident in children aged 5-9 years.

##### RESEARCH QUESTION 2: Daily versus Intermittent Supplementation

Three SRMAs compared daily and intermittent prophylaxis (one, two or three times a week on non-consecutive days) in pregnant women(78,82,86), one in WRA(75),and one in children(66)In addition, ICMR has carried out an SRMA comparing daily versus intermittent regimens in all age groups (Definitions and durations for all mentioned in Supplementary Table 05)

### Haemoglobin level

#### Children, adolescents, and WRA

No significant differences in haemoglobin were reported between daily and intermittent regimens in children 6-59 months: children and adolescents <12 years: MD -0.60 g/L (95% CI: -1.54, 0.35 g/L; high quality)(67), Women of reproductive age: MD 0.43 g/L (95% CI: -1.44, 2.31 g/L; high quality)(76)

#### Pregnant women

Findings were mixed. One high-quality review found no difference in haemoglobin levels between daily and intermittent supplementation (SMD 0.51, 95% CI: -0.23, 1.24) or weekly supplementation (SMD 0.89, 95% CI: 0.07, 1.84)(86). Peña-Rosas et al. reported lower risk of haemoconcentration (Hb >130 g/L) with intermittent supplementation (RR 0.53, 95% CI: 0.38, 0.74; high quality)(87).

### Anaemia Risk

Data comparing anaemia risk between intermittent and daily supplementation are limited.

#### Children <12 years

Higher anaemia risk with intermittent supplementation (RR 1.23, 95% CI: 1.04, 1.47; high quality)(67)

#### WRA

No significant difference (RR 1.09, 95% CI: 0.93, 1.29; high quality)(76)

#### Pregnant women

No significant difference (RR 1.20, 95% CI: 0.78, 1.83; high quality)(82); RR 1.22 (95% CI: 0.84, 1.80; high quality)(87)

### Ferritin Levels

Daily supplementation was associated with higher ferritin levels in pregnant women (SMD 0.85; 95% CI: 0.15–1.54; p = 0.02; high quality)(86) and in women of reproductive age (MD 6.07 ng/mL; 95% CI: 1.48–10.66 ng/mL; p < 0.001; high quality)(76). No significant difference was observed in children under 12 years (MD –4.19 ng/mL; 95% CI: –9.42 to 1.05 ng/mL; high quality) (67).

### Iron Deficiency and Iron Deficiency Anaemia

#### Children <12 years

Intermittent supplementation increased ID risk (RR 4.00, 95% CI: 1.23, 13.05; high quality)(67)

#### Women of Reproductive Age

No significant difference in ID risk (RR 4.30, 95% CI: 0.56, 33.20) (76)

#### Pregnant women

No significant difference in IDA at term (RR 0.71, 95% CI: 0.08, 6.63; high quality)(87)

Data were unavailable for adolescents or preschool children.

### Adverse Birth outcomes

No differences between the intermittent and daily group was reported for low birthweight (RR 0.82; 95% CI 0.55 to 1.22 ), infant birthweight (MD 5.13 g; 95% CI -29.46, 39.72g), premature birth (RR 1.03; 95% CI 0.76 to 1.39)(87).

### Adverse effects and adherence

No significant differences in side effects were observed among children under 12 years (RR 0.60, 95% CI: 0.19,1.87;*high quality*)(67).Among pregnant women, high quality reviews reported that intermittent supplemented group reported lower gastric side effects like nausea(AOR 3.56, 95% CI: 2.23,5.69, p < 0.001), diarrhoea(AOR 5.40, 95% CI: 1.90,15.33, p = 0.002), vomiting (AOR: 3.22, 95% CI: 0.94,10.95, p = 0.06)constipation(AOR 1.95, 95% CI: 1.21,3.14, p = 0.006)(86).De Regil et al.reported no difference in adherence between two groups(67).

### Regional data from India and South Asia

#### Haemoglobin

Regional data from India and South Asia shows that daily as compared to intermittent supplementation significantly improved haemoglobin levels in 5–9 years children (MD of 4.93 g/L 95% CI: 1.13g/L, 8.73g/L; p = 0.01, *4 studies*) (28,31,45)(58) and pregnant women(MD was 3.11 g/L 95% CI: 1.00g/L,5.22g/L; p< 0.001;*9 studies*)(23,31,33,36,41,42,45,49,51). No studies on daily versus intermittent prophylaxis children (6–59 months), adolescents, or WRA could be identified. (Most studies compared daily versus intermittent treatment regimen)

#### Ferritin

The effect on serum ferritin among 5-9 y old children was negligible (MD 0.13 ng/mL; 95% CI: -4.32ng/mL, 4.57ng/mL;*2 studies*)(29,46), while it was significantly higher with daily supplementation among pregnant women (MD 12.74 ng/mL; 95% CI: 5.61ng/mL, 19.88ng/mL; p = 0.001; *2 studies*)(23,41). (Supplementary figure 6-9). No studies were identified among non-anaemic, healthy children (6–59 months), adolescents, or WRA

### Summary of evidence for Research Question 2 (Efficacy of daily versus intermittent iron/ iron folic acid supplementation)

To summarize, high-quality systematic reviews found no significant difference in haemoglobin levels between daily and intermittent iron supplementation among children aged 6–59 months, older children, adolescents, or women of reproductive age(67,76) . For pregnant women, findings were mixed: one review reported no difference between regimens (86). Another review found a lower risk of haemo-concentration with intermittent dosing(87).

Anaemia risk: Data were limited. Evidence from a single high-quality study reported higher anaemia risk among children under 12 years receiving intermittent supplementation(67), while no significant difference was observed among WRA(76) or pregnant women(87).

Ferritin levels: Across high-quality studies, daily supplementation resulted in higher ferritin levels in WRA(76) and pregnant women(86), while no significant difference was found in children under 12 years(67).

Iron deficiency and IDA: Across high-quality studies, intermittent supplementation increased ID risk in children <12 years(67), but no significant difference was observed in WRA(76). For pregnant women, no significant difference in IDA at term was reported(87). Data were unavailable for adolescents or preschool children.

Adverse Birth outcomes and adherence: Across high-quality studies, intermittent and daily supplementation showed no differences in adverse birth outcomes (low birth weight, preterm birth, stillbirth) or adherence(67,87). However, among pregnant women, intermittent supplementation was associated with fewer gastrointestinal side effects and better tolerability(86).

Regional evidence: Limited data from India and South Asia suggest that daily supplementation produces greater haemoglobin gains and higher ferritin concentrations in pregnant women, and higher haemoglobin levels—but no difference in ferritin—in children aged 5-9 years. No comparative data were available for other age groups.

### RESEARCH QUESTION 3: Adverse effects of iron/IFA supplementation

Evidence on the adverse effects of iron prophylaxis across target groups, as documented in SRMAs, is summarized in Supplementary Table 6 and briefly discussed below. However, there were insufficient studies from India and South Asia.

#### Gastro-intestinal side effects

There was limited evidence comparing side effects of supplementation in non-anaemia/non-iron deficient, hence we also compared the adverse effects in supplemented and non-supplemented groups.

#### Children

In children aged 4–59 months, Ghanchi et al.(88) reported no significant increase in diarrhea but a higher prevalence of bloody diarrhea in iron-replete individuals. Due to the paucity of evidence comparing adverse effects of iron supplementation in non–iron-deficient versus iron-deficient children, we also compared adverse effects between supplemented and non-supplemented groups from Question 1 (irrespective of iron status). Pasricha et al. found a higher risk of vomiting in children aged 4–23 months (RR 1.38; 95% CI: 1.10, 1.73; p=0.006; *high quality*) (71).

#### WRA

Diarrhea (RR 2.13; 95% CI: 1.10, 4.11; p=0.31), constipation (RR 2.07; 95% CI: 1.35, 3.17; p=0.77), and abdominal pain (RR 1.55; 95% CI: 0.99, 2.41)(75) were more common in supplemented group.

#### Pregnant women

An SRMA by Watt et al. (2025) among 4,492 non-anaemic pregnant women found that prophylactic oral iron supplementation increased haemoglobin and ferritin levels and reduced anaemia risk by 50%, with no significant effects on adverse birth outcomes however there was insufficient evidence for any conclusion regarding adverse effect of supplementation in non-anaemic women (80).

### Impact on Growth Outcomes

No SRMA compared growth outcomes between anemic/iron-deficient individuals and non-anemic/non-iron-deficient individuals supplemented with iron. Therefore, we compiled growth outcomes from other SRMAs. Pasricha et al. found no difference in final length, weight, or related Z-scores between iron-supplemented children (4–23 months) and controls. However, when measured as change from baseline, iron supplementation was associated with reduced length gain (SMD -0.83; 95% CI: -1.53, -0.12; p=0.02) and weight gain (SMD -1.12; 95% CI: - 1.91, -0.33; p=0.0005). Children receiving iron for more than three months had lower weight gain (SMD -2.39; 95% CI: -4.37, -0.41; p=0.02;*high quality*)(71). Sachdev et al. (2006) also reported a small negative effect on linear growth with long-term supplementation (>6 months) (SMD -0.13; 95% CI: -0.24, -0.01) and in children from developed countries (SMD -0.27; 95% CI: -0.49, -0.05; p=0.018; *moderate quality*)(73). However, three reviews all high-moderate quality (71,73,74) found no impact on growth or anthropometry in preschool children, suggesting no overall adverse effect of iron supplementation on growth outcomes.

### Adverse Birth outcomes

Two high quality SRMAs compared birth outcomes in non-anemic/ iron-replete women supplemented versus non anemic controls who were not supplemented. Watt et al. found no significant difference in cesarean rates (OR 1.07; 95% CI: 0.88, 1.29) or preterm births (OR 0.82; 95% CI: 0.58, 1.17)(80) in non-anemic women given iron/IFA versus those not supplemented. Hansen et al. reported lower LBW (RR 0.30; 95% CI: 0.13, 0.68) and SGA (RR 0.39; 95% CI: 0.17, 0.86)(81) in iron-replete women receiving iron versus controls.

### Oxidative Stress, gut microbiota and gestational diabetes milletus

No SR/SRMA on oral IFA supplementation and oxidative stress were identified. Therefore, findings from observational studies and RCTs are summarized. One study found iron supplementation increased oxidative stress and inflammation (hsCRP) in non-anemic women, while CRP levels decreased in anemic women without altering oxidative stress markers. Baseline oxidative stress and inflammation were higher in anemic than non-anemic pregnant women (89).Shastri et al., observed that iron supplementation in healthy, nonanemic pregnant women increased oxidative stress(424·7 (SD 163·7) versus 532·9 (SD 144·7) pmol p-nitrophenol formed/min per ml plasma, p=0·002)(90). These findings suggest that the effects of iron supplementation on oxidative stress depend on baseline iron status, benefiting those with compromised iron status while increasing oxidative stress in individuals with adequate iron status.

Ghanchi et al. reviewed 19 studies regarding gut microbiota, of which 12 found no difference in diarrhea between supplemented and non-supplemented groups. The remaining studies reported an increase in diarrhea in the supplemented group(88). An SRMA on iron supplementation or fortification in infants found a 6.37% reduction in bifidobacteria populations (95% CI: 10.16–25.8%)(91).

Two SRMAs examined the association of iron with gestational diabetes mellitus (GDM)(92,93). Kataria et al. reported that women with GDM had higher iron biomarker levels compared to those without GDM –SMD 0.25 (95% CI: 0.001, 0.50) for iron, 1.54 ng/mL (95% CI: 0.56,2.53ng/mL) for ferritin, 1.05% (95% CI: 0.02,2.08) for transferrin saturation, and 8.1 g/L (95% CI: 4.0,12.2g/L) for hemoglobin. The adjusted odds ratios (OR) for GDM were 1.58 (95% CI: 1.20,2.08) for ferritin, 1.30g/L (95% CI: 1.01,1.67) for Hb, and 1.48 (95% CI: 1.29,1.69) for dietary heme iron intake. However, the review cautioned that the high heterogeneity of analyses limited the interpretability of findings(92). Similarly, Miranda et al. could not provide conclusive evidence on this association (93).

To summarise short-term studies reported no significant adverse effects beyond gastrointestinal intolerance, which was more common with daily compared to intermittent supplementation. No serious adverse events were attributed to prophylactic iron supplementation. However, a critical knowledge gap exists regarding long-term safety in non-iron-deficient individuals. Emerging evidence suggests potential concerns including oxidative stress, gut microbiome alterations, and possible associations with gestational diabetes, warranting further investigation

### Regional data from India and South Asia

Only ten trials (54, 53, 23, 50, 17, 38, 18, 40, 56, 31) reported side effects across age groups, which limited the ability to conduct detailed subgroup analyses. Four trials included pregnant women; three of these reported that daily supplementation was associated with more adverse effects than intermittent dosing. Gastrointestinal discomfort was reported in three daily and two intermittent studies overall. In adolescents, one study documented constipation, nausea, vomiting, epigastric discomfort and rash in the daily group. Among children 5–9 years, one study reported constipation and nausea/vomiting, while in children 6–59 months daily supplementation was associated with respiratory symptoms (three studies) and diarrhoea/dysentery (two studies). One study reported that weekly supplementation resulted in diarrhoea, nausea/vomiting and epigastric discomfort among women of reproductive age.

## DISCUSSION

This comprehensive review of 21 systematic reviews (17 high-quality, 4 moderate-quality) and meta-analysis of 44 regional studies from India and South Asia provides robust evidence supporting iron/IFA prophylaxis for anaemia control across all age groups in India.

Prophylactic iron/IFA supplementation consistently improved haematological outcomes, with observed haemoglobin increases of 4–9 g/L, ferritin increments of 7–28 ng/mL, and anaemia risk reductions ranging from 23–70%. There were variations in the in the extent of improvement across age groups . The pattern of risk reduction provides insights into the relative contribution of iron deficiency to anaemia across age groups. Among pregnant women, a 70% reduction in anaemia risk and 77% reduction in IDA risk were observed, underscoring iron deficiency as the predominant cause. Similarly, in WRA, anaemia (35–61%) and ID (38–50%) risk reductions were closely aligned. In children under five years, IDA risk reduction (68–86%) exceeded overall anaemia risk reduction (23–45%), suggesting iron deficiency as a major but not sole contributor. In older children and adolescents, despite high IDA risk reduction (91%), anaemia risk reduced by only approximately 50%, implying that nearly half of anaemia cases in these groups may be attributable to non-iron causes.

Even under controlled trial conditions, the haemoglobin and ferritin gains with IFA supplementation were modest, indicating that when implemented at scale, the population-level impact is likely to be smaller due to, infection burden, and non-iron causes of anaemia. These findings underscore that IFA supplementation while effective, should be part of a comprehensive anaemia control strategy that includes various complementary strategies.

For all age groups except pregnant women, haematological outcomes were comparable between daily and intermittent regimens. Among pregnant women, daily supplementation was superior for haemoglobin and ferritin levels, though intermittent regimens had fewer gastrointestinal side effects. The AMB program provides weekly IFA prophylaxis to all age groups except pregnant women ( biweekly to children 6-59 months) , differing from the WHO recommendation, which advises daily prophylaxis where prevalence of anaemia is >40%(10). Evidence from this review indicates that intermittent (primarily weekly) and daily iron supplementation result in comparable Hb increment across most age groups, except pregnant women and children. Regional analysis favoured daily supplementation for children 5-9 years of age and global evidence indicates that intermittent supplementation was associated with an increased risk of anaemia and iron deficiency (ID) among children under 12 years globally(67), despite no overall difference in mean ferritin levels. Among WRA, anaemia and ID risks were similar between regimens, although daily supplementation more effectively improved ferritin levels. In pregnant women, no differences were found in birth outcomes, anaemia risk, or IDA between daily and intermittent (mostly once-weekly) supplementation. However, haematological findings were mixed. The review by Banerjee et al. (86) reported no difference in Hb , the ICMR unpublished review found higher Hb and ferritin with daily dosing. A recent SRMA by Chillo et al. (94) reported lower Hb with intermittent supplementation (MD −2.4 g/L; 95% CI: −3.5, −1.2). Regional data further show better Hb levels and higher ferritin levels in pregnant women with daily dosing. Across SRMAs, fewer gastrointestinal side effects were reported with intermittent prophylaxis during pregnancy.

Based on this evidence, daily prophylaxis should be recommended for all pregnant women, as pregnancy is the only window to build iron stores, given the high prevalence of anaemia in the postpartum period when IFA adherence drops significantly. Intermittent supplementation may, however, serve as a feasible alternative in cases where gastrointestinal intolerance limits daily compliance. Currently, the AMB programme does not include alternate-day or intermittent options for pregnant women—an approach that could be considered for future inclusion. Both global and Indian evidence indicate a potential benefit of daily supplementation among school-age children, though data remain limited in this age group. This highlights the need for further research to understand the optimal dosing frequency for children and adolescents in high-anaemia settings.

Data on the long-term safety of IFA supplementation, particularly in non-iron deficient individuals, is limited. The effects of iron supplementation on oxidative stress and GDM remain unclear, requiring further research. Given the limited data on long-term adverse effects and the observation that hemoglobin improvements and cognitive improvements are greatest in individuals anemic at baseline, targeted approaches could minimize unnecessary supplementation, reduce risks like gastrointestinal intolerance and oxidative stress, and maximize benefits. In this context, ICMR’s efforts to develop indigenous point-of-care (POC) devices for ferritin measurement could significantly improve the precision of iron deficiency diagnosis and treatment (ICMR EOI for POC Devices).

Our findings align with previous global systematic reviews demonstrating efficacy of iron supplementation for improving haematological outcomes. However, this review makes several unique contributions. The meta-analysis of 44 studies from India and South Asia confirms that global findings apply to regional populations, with comparable effect sizes. This is critical for policy decisions in the Indian context .Our synthesis demonstrates that weekly supplementation is effective even where anaemia prevalence is high, providing evidence to support the AMB approach. Our findings support offering an intermittent iron supplementation regimen for pregnant women in India who experience gastrointestinal intolerance with daily dosing, an option not presently included in the national programme. While evidence remains limited, our synthesis highlights the lack of growth benefits and mixed cognitive findings, tempering expectations about any broader developmental impacts of iron supplementation.

### Strengths and Limitations

This review has several notable strengths. First, it provides comprehensive synthesis of evidence across multiple age groups, encompassing 21 systematic reviews and 44 regional studies. Second, the focus on region-specific data from India and South Asia provides valuable insights for programme implementation in high-burden settings. Third, quality assessment using AMSTAR ensures standardized evaluation of review quality, with the majority rated as high quality. Fourth, the inclusion of data on both haematological and non-haematological outcomes, as well as adverse effects and regimen comparisons, provides a holistic evidence base for policy decisions. Finally, direct relevance to AMB programme design makes this review immediately actionable for policy and practice.

Several limitations must be acknowledged. First, we did not account for heterogeneity within the included systematic reviews, which could influence the reliability of pooled estimates. The high I² values observed in our meta-analyses indicate substantial heterogeneity that was only partially explained by sample size and intervention duration. Second, potential overlap of primary studies across reviews poses a risk of data duplication, potentially inflating the strength of evidence, though this is an inherent limitation of reviews of reviews. Third, our search was limited to PubMed and Cochrane databases, potentially excluding relevant studies from other sources, though these are the primary databases for systematic reviews. Fourth, the evidence base for certain outcomes remains limited, particularly for non-haematological effects, long-term safety, and the impact of supplementation in specific subgroups such as WRA. Finally, we found limited studies from India and South Asia both within global SRMAs and through additional searches, limiting the precision of region-specific estimates for some age groups.

## Conclusion

This review shows that iron /iron folic acid prophylaxis is helpful in reducing anaemia and IDA, IDacross all beneficiary groups. The findings supports daily supplementation for pregnant women and weekly supplementation for other groups. There is no conclusive evidence of adverse effects at prophylactic doses, targeted supplementation for iron-deficient individuals may enhance both the safety and effectiveness of iron prophylaxis. Future research should focus on long-term safety, non-hematological outcomes, and strategies to improve adherence and program delivery, ensuring that the benefits of IFA supplementation are fully realized in real-world settings.

## Supporting information

PRISMA checklist

## AUTHORS’ CONTRIBUTION

RM led all aspects of the review data curation (title and abstract screening, full-text screening, and data extraction), formal analysis, project administration, supervision, validation, and writing (original draft, review, and editing).BK conceptualised the review and was involved in project administration, supervision and manuscript writing and editing. SM, TL VK, RC, RP, MKN,MJ,PT and AJ were involved in data curation (full-text screening and data extraction), validation, and writing (review and editing).RC and RP also were involved in manuscript writing and editing.BK, MKN and PR, RC, MJ and RM also critically reviewed the final manuscript. SM, RM and VK primarily extracted the data and perform statistical analysis. All authors critically reviewed the manuscript and approved the final manuscript for submission.

## DECLARATION OF INTEREST

All authors declare no competing interests.

## DATA AVAILABILITY

All data in this review were from publicly available systematic review and meta-analysis and RCTs

## ACKNOWLEDGEMENTS

Ms Preetu Mishra, UNICEF India Country Office; Ms Richa Singh Pandey, UNICEF India Country Office; Dr NK Arora, The INCLEN trust, New Delhi; Dr Aakriti Gupta, NITI Aayog; Dr Zoya Ali Rizvi, MOHFW.

## FUNDING

No funding was received for this review

---

**Table 1:**
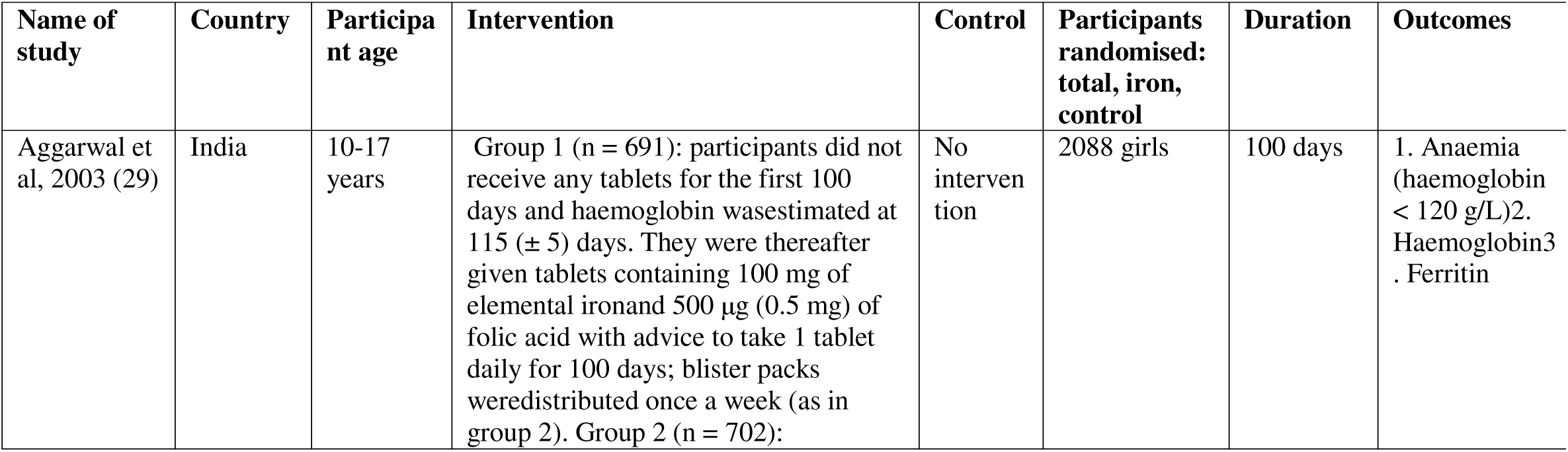
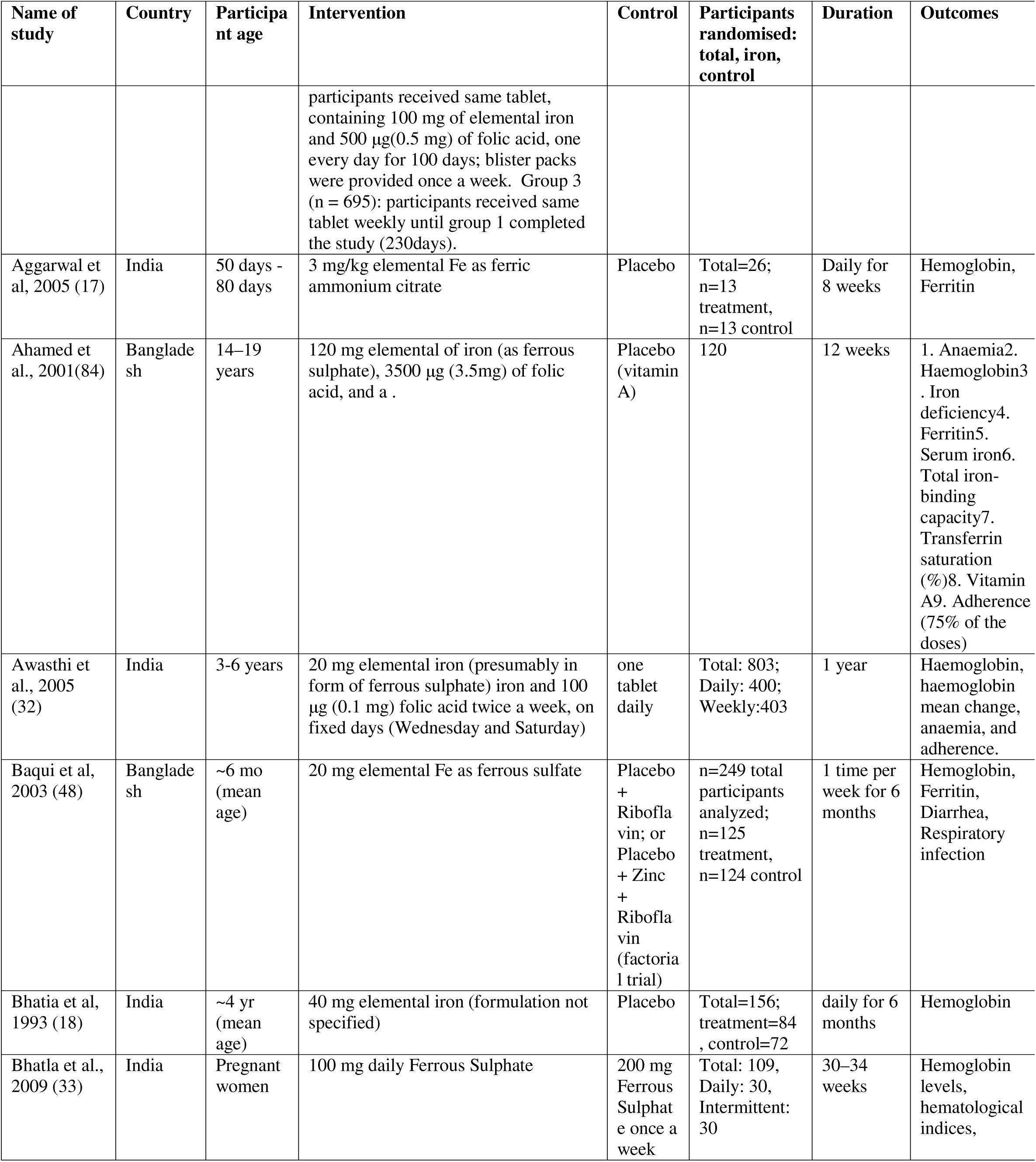
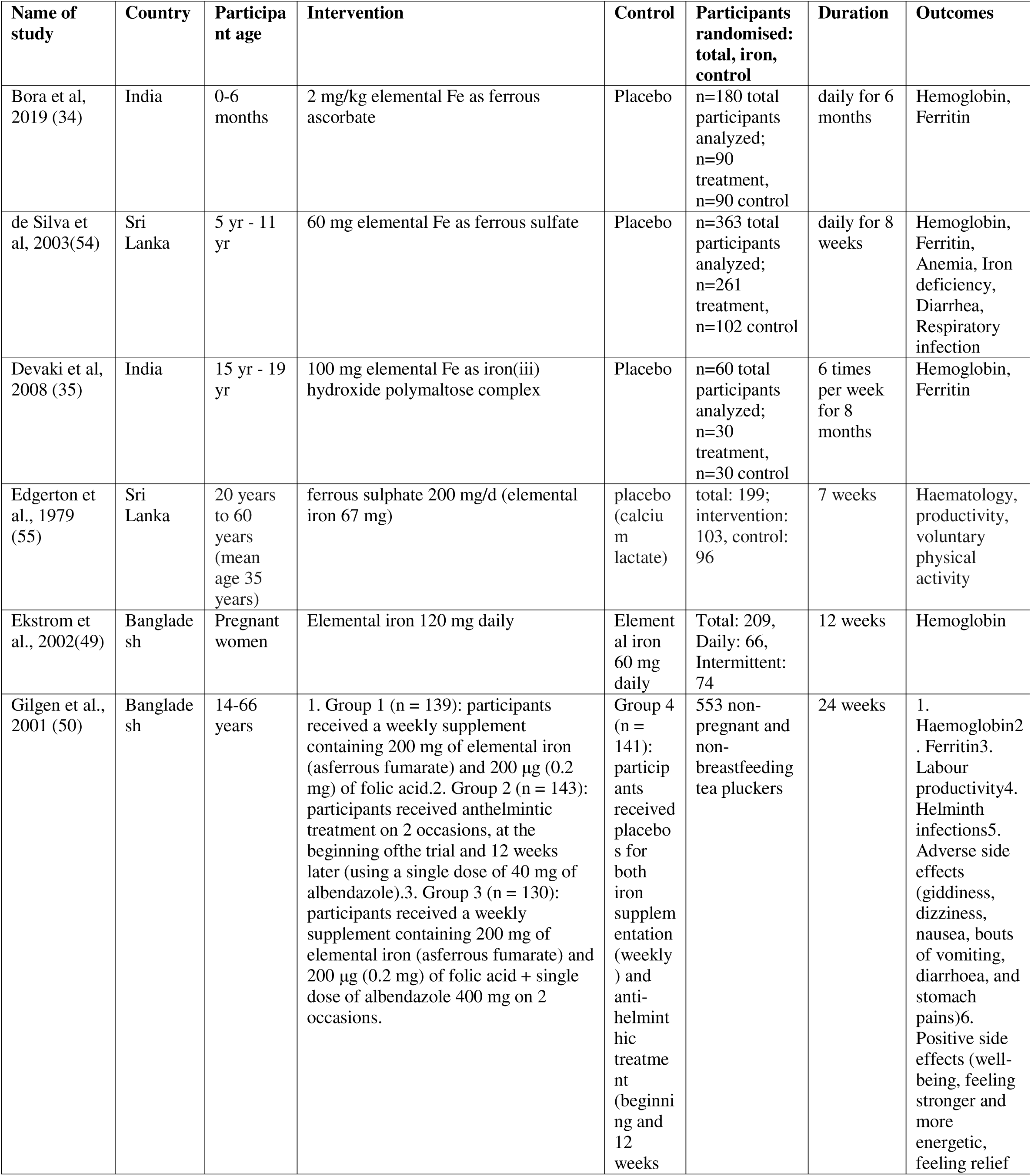
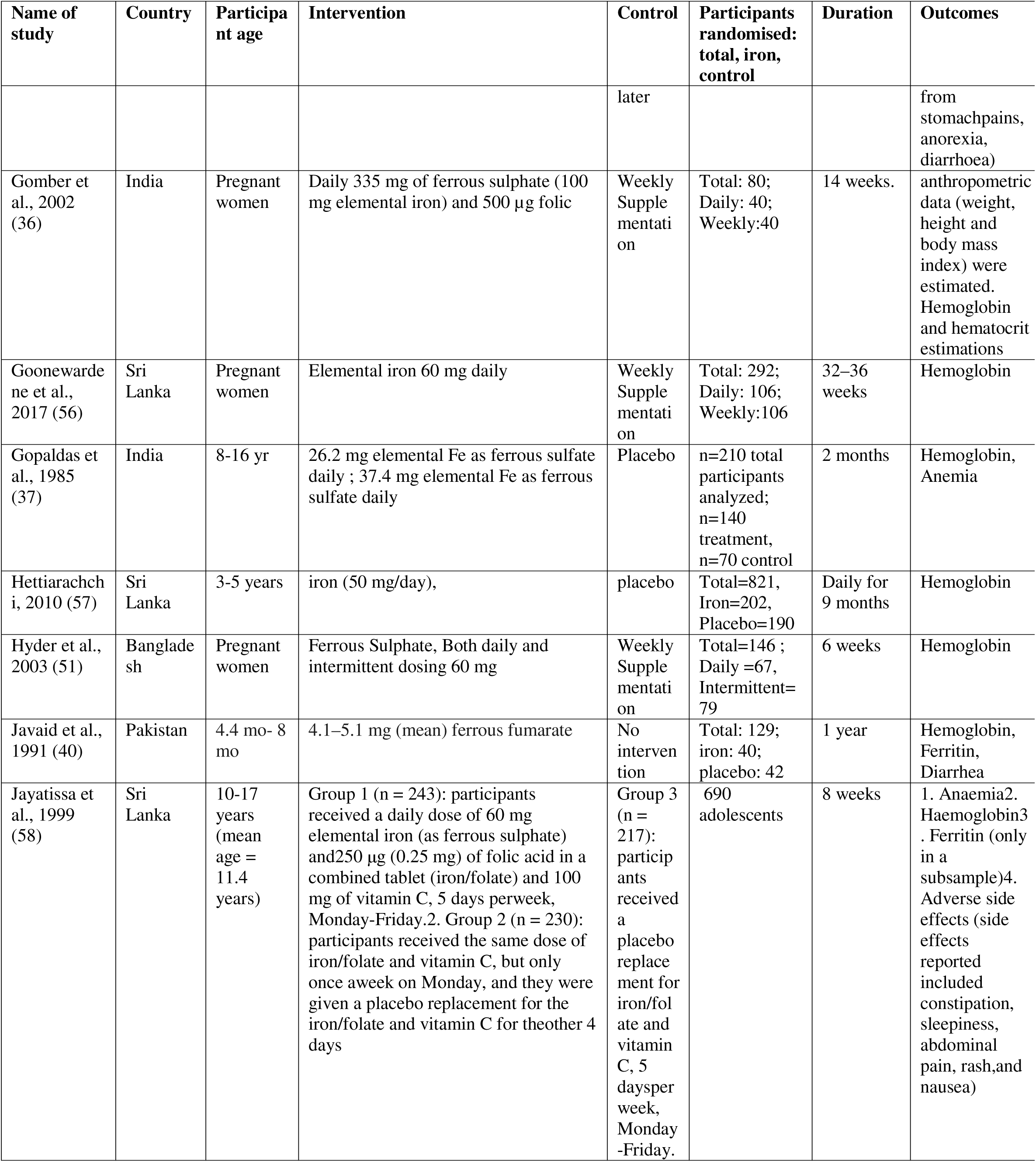
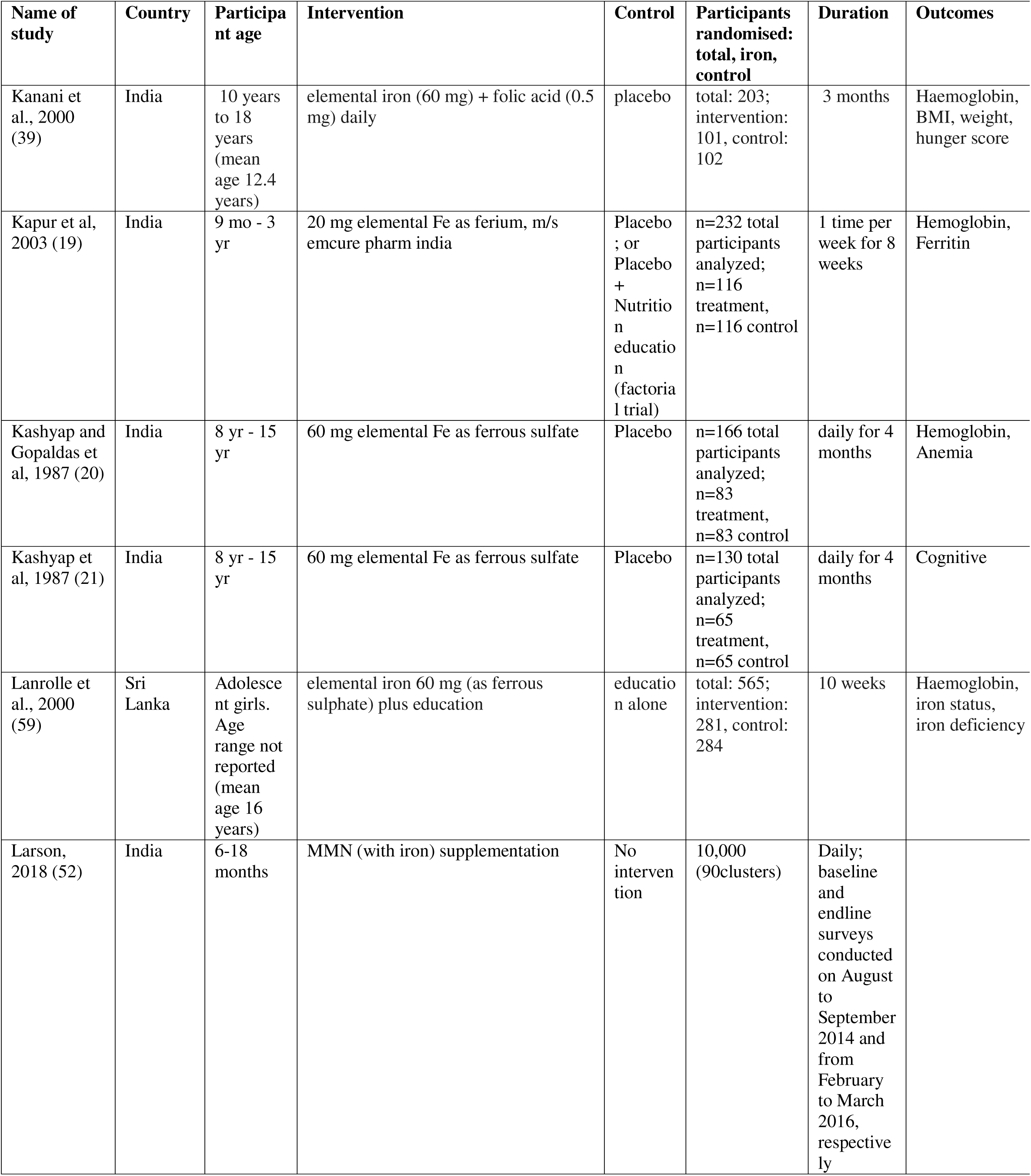
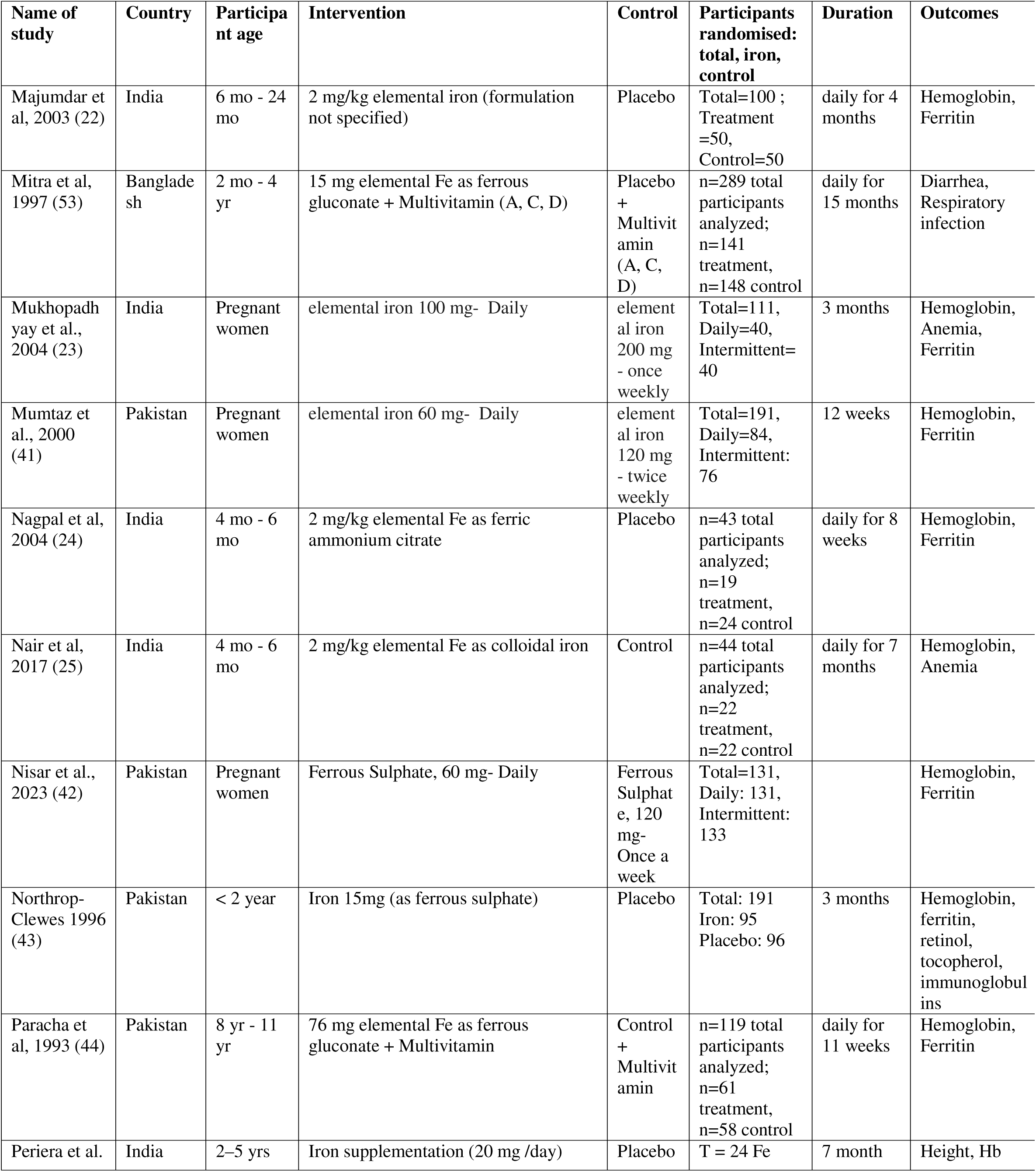
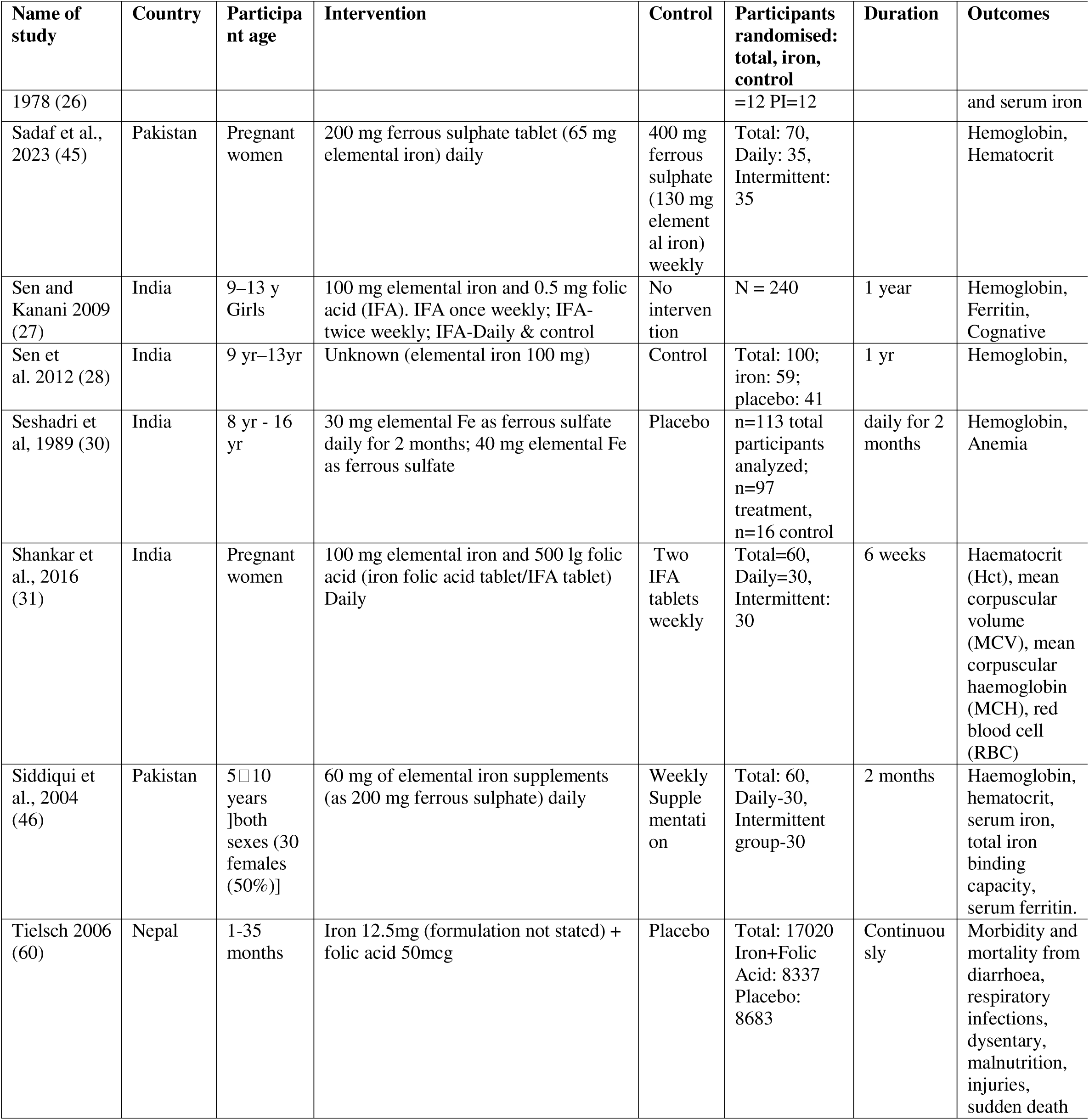
Study characteristics of included South Asian Studies.

**Figure 1:**
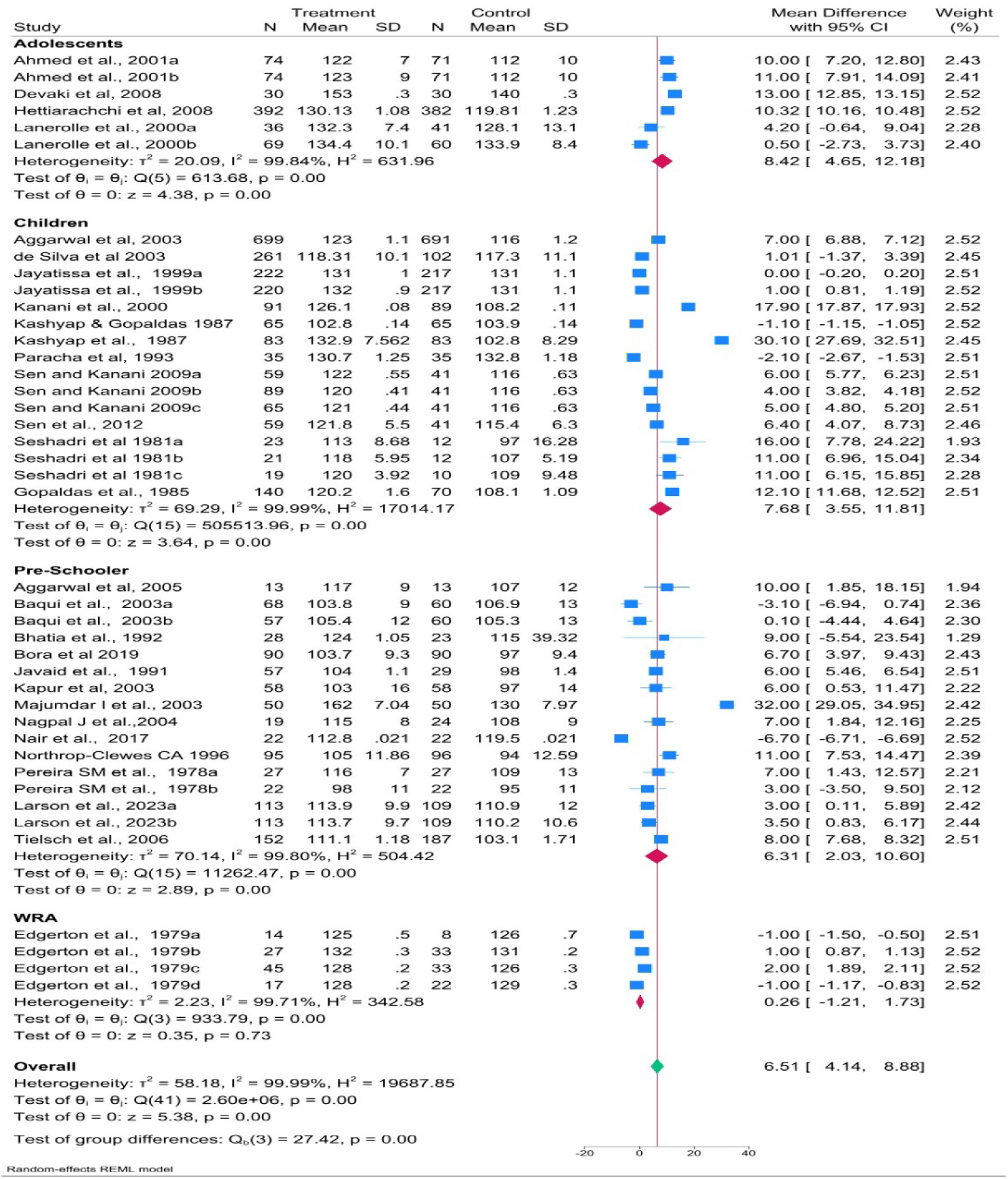
Effect of Iron prophylaxis on Hemoglobin Levels (g/L) in Indian and South Asian Studies across Different Age Groups.

**Figure 2:**
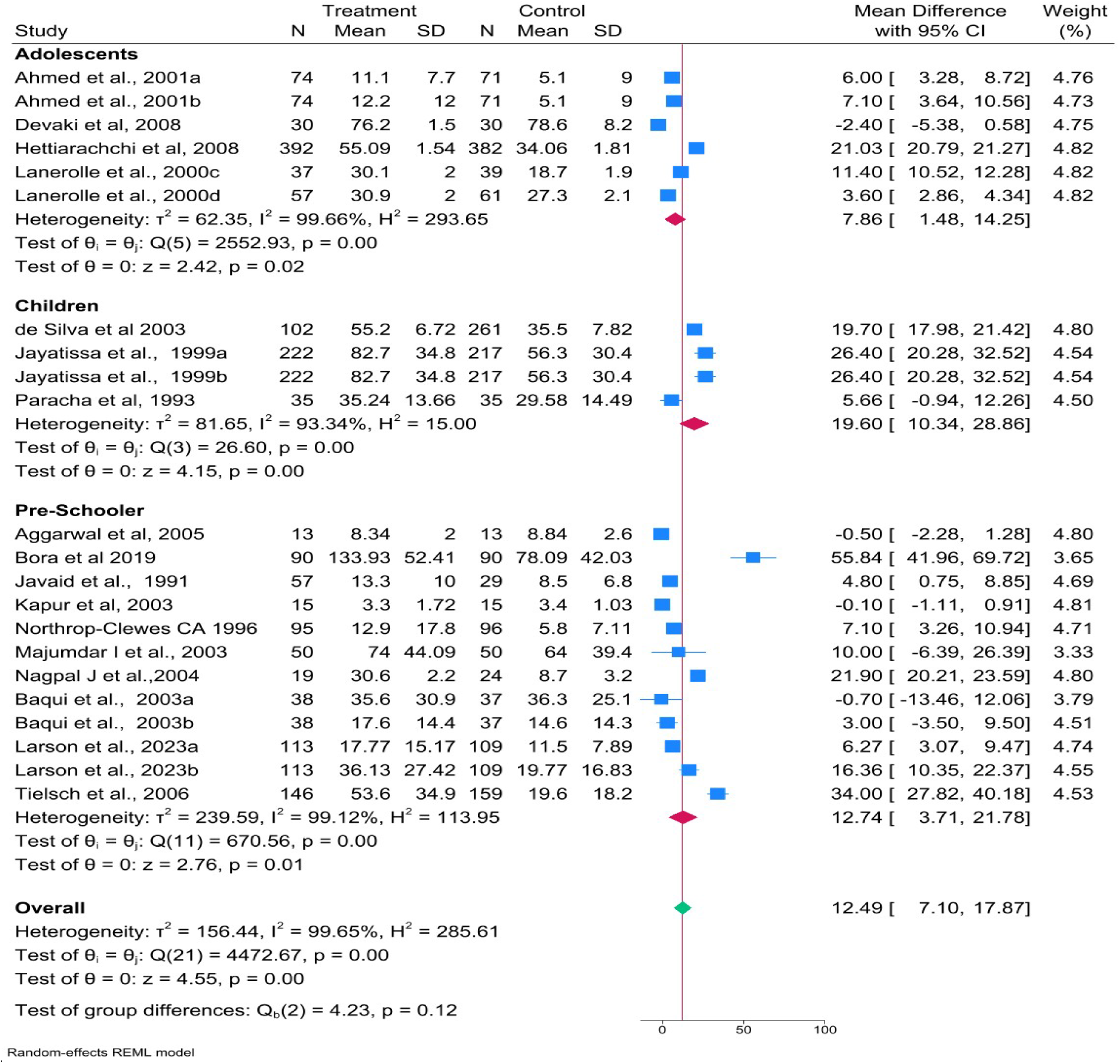
Effect of Iron prophylaxis on Ferritin (ng/mL) Levels in Indian and South Asian Studies across Different Age Groups.

**Figure 3:**
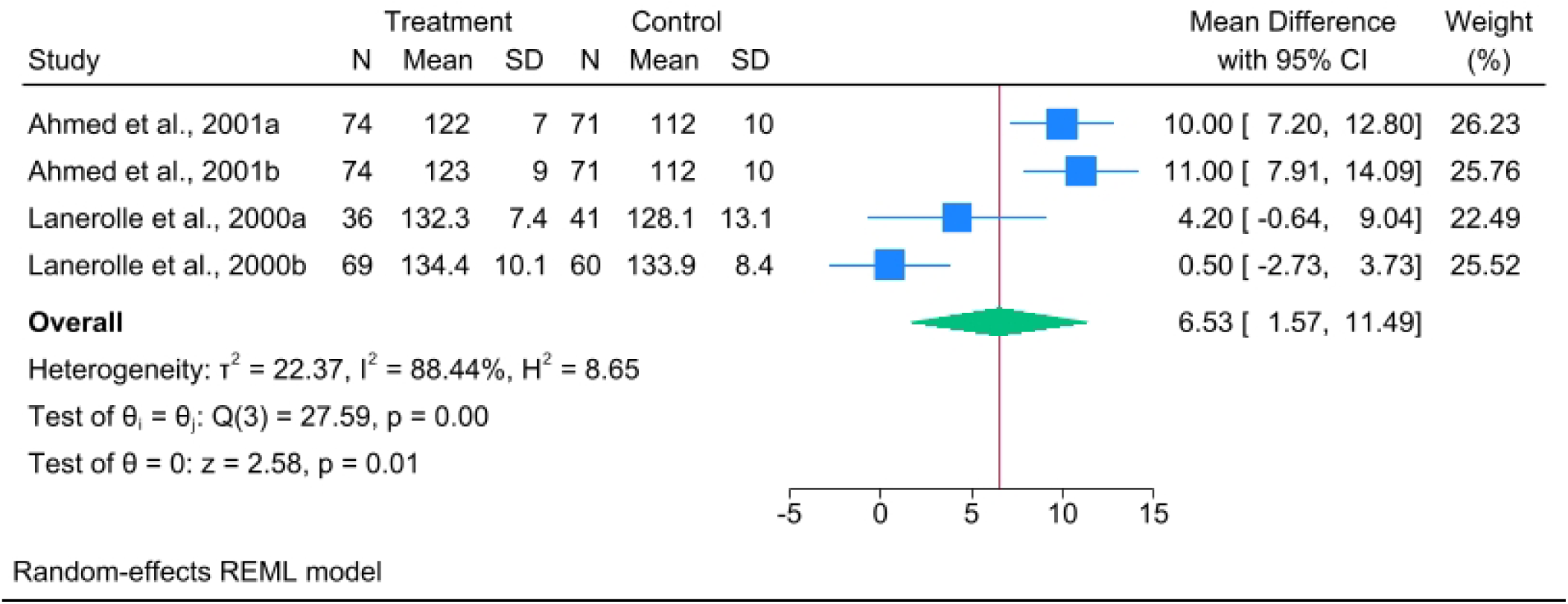
Effect of Iron prophylaxis on Hemoglobin Levels (g/L) in Indian and South Asian Studies for adolescent girls.

**Figure 4:**
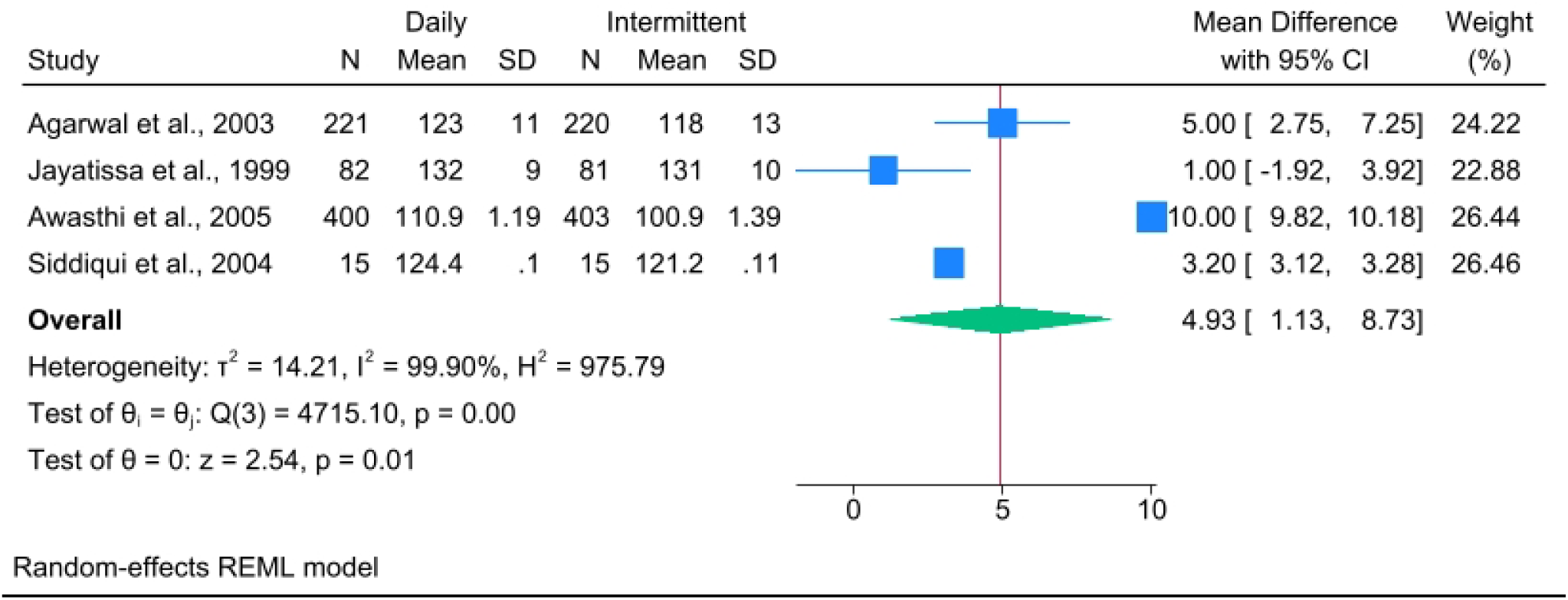
Effect of Daily vs Intermittent iron prophylaxis on Haemoglobin concentration (g/L) of Children (Indian and South Asian studies)

**Figure 5:**
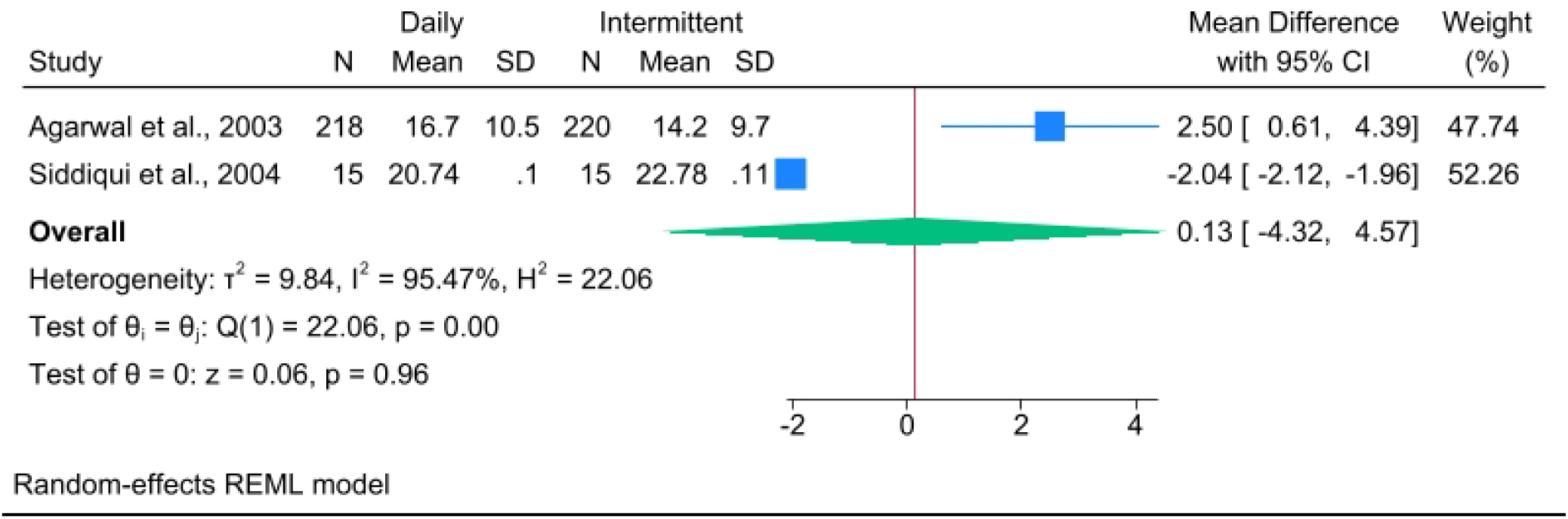
Effect of Daily vs Intermittent Interventions on Ferritin levels (ng/mL) in Children (Indian and South Asian studies) (Indian and South Asian <u>studies)</u>.

**Figure 6:**
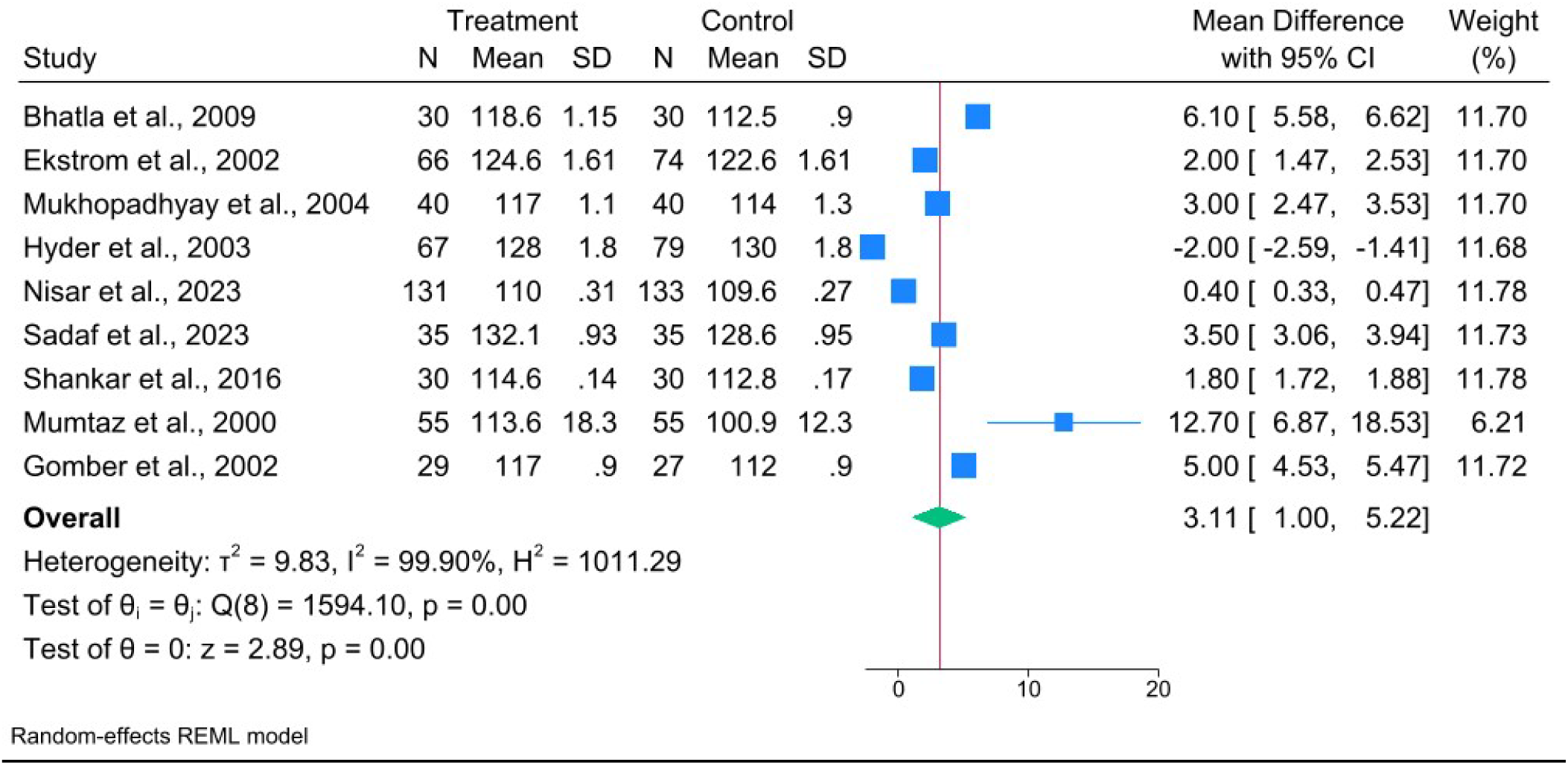
Effect of Daily vs Intermittent Iron Intervention on haemoglobin levels (g/L) in Pregnant women (Indian and South Asian <u>studies</u>)

**Figure 7:**
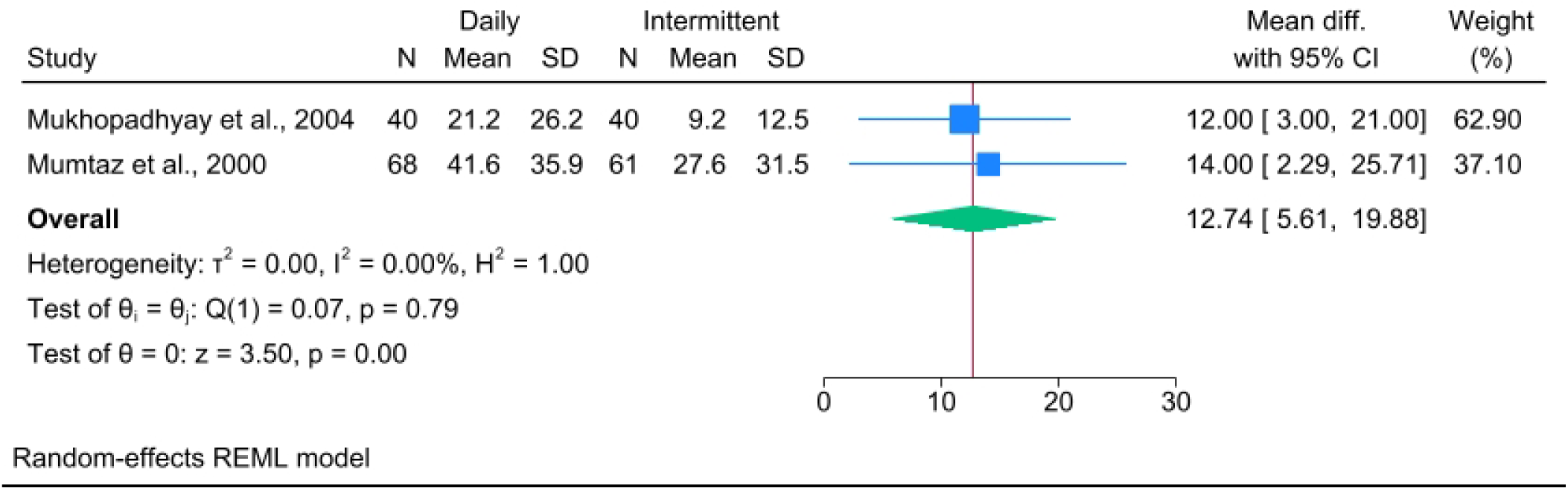
Effect of Daily vs Intermittent Interventions on Ferritin levels (µg/L) of Pregnant women (Indian and South Asian studies)

**Figure 8:**
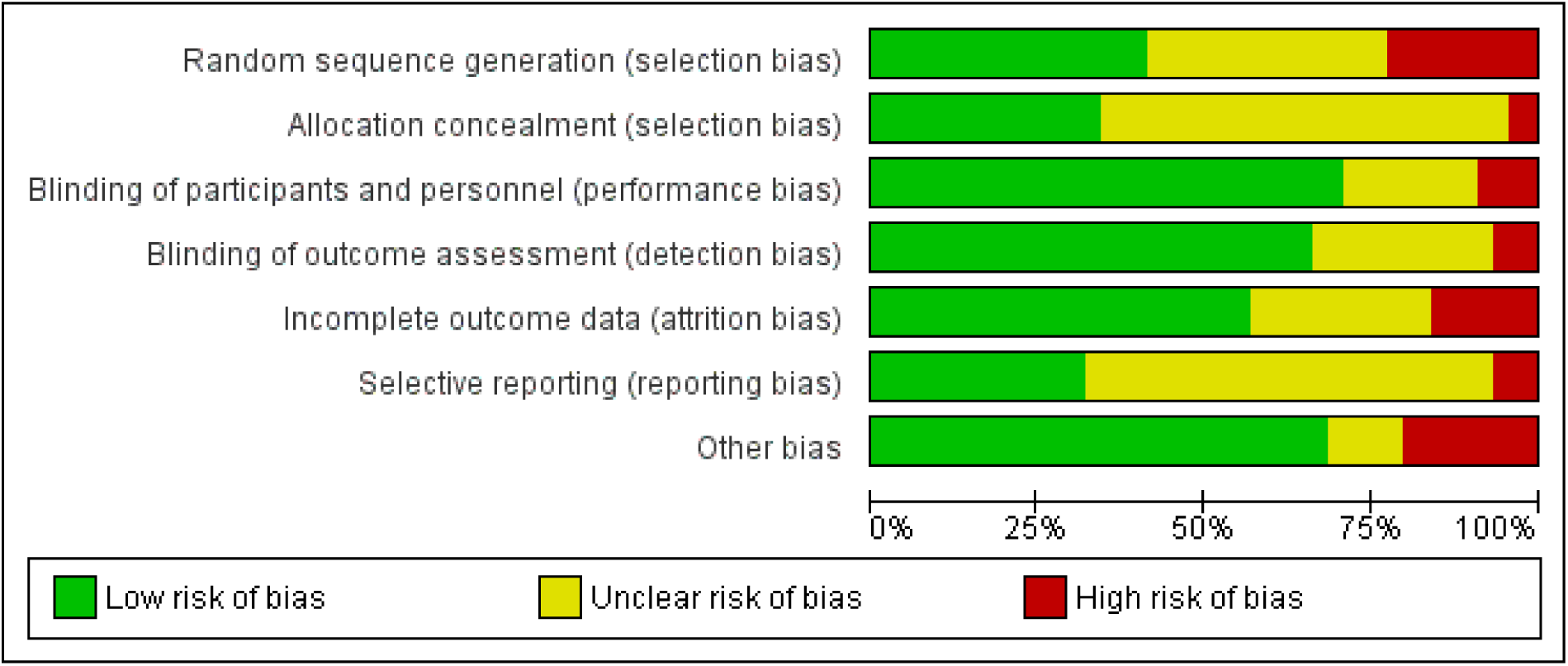
Risk of Bias of Indian and South Asian studies.

**Figure 9:**
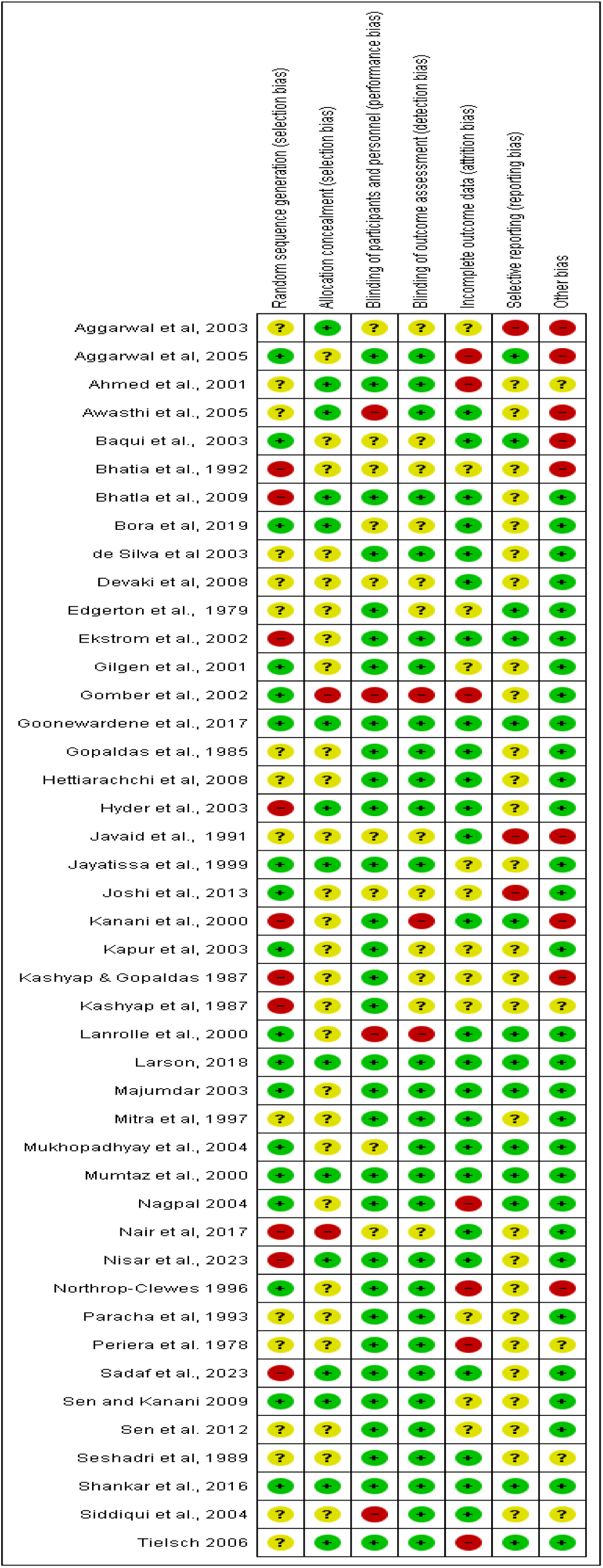
Risk of Bias Summary of Indian and South Asian studies.

## Notes

### Competing Interest Statement

The authors have declared no competing interest.

### Funding Statement

This study did not receive any funding

