## Supplemental Tables for "A Comprehensive Review of Iron Prophylaxis in the National Anemia Control Programme in India (Anemia-Mukt Bharat)"

Supplementary [Table no 3: AMSTAR-I ratings for each systematic review	5](#_Toc213429487)

Supplementary [Table 4: Effectiveness of iron supplementation in various age groups	0](#_Toc213429488)

Supplementary [Table 5: Daily versus intermittent iron prophylaxis on haemoglobin ferritin levels in various age groups	8](#_Toc213429489)

Supplementary [Table 6: Adverse effects with iron supplementation in various age groups	10](#_Toc213429490)

### Supplementary Table 1: Current national and World Health Organisation (WHO) recommendations for iron prophylaxis in various age groups

| **Age group** | **Dosage and regimes (AMB, India)** | **WHO Recommendations** |
| --- | --- | --- |
| 6 – 59 months of age | Biweekly, 1 ml Iron and Folic Acid syrup  Each ml of Iron and Folic Acid syrup containing 20 mg elemental Iron + 100 mcg of Folic Acid | Daily iron supplementation (10-12.5 mg elemental iron) for three consecutive months a year is recommended as a public health intervention in infants and young children aged 6–23 months, living in settings where the prevalence of anaemia is 40% or higher in this age group*, for preventing iron deficiency and anaemia. (10)  Daily iron supplementation(30 mg elemental iron) for three consecutive months a year is recommended as a public health intervention in preschool-age children aged 24–59 months, living in settings where the prevalence of anaemia in infants and young children is 40% or higher, for increasing haemoglobin concentrations and improving iron status. (10)  If the prevalence of anaemia is 20–40%, intermittent regimens of iron supplementation can be considered. |
| 5- 10 years children | Weekly, 1 Iron and Folic Acid tablet  Each tablet containing 45 mg elemental Iron + 400 mcg Folic Acid, sugar-coated, pink colour | Daily iron supplementation (30-60 mg elemental iron) for three consecutive months a year living in settings where the prevalence of anaemia is 40% or higher in this age group (10)  If the prevalence of anaemia is 20–40%, intermittent regimens of iron supplementation can be considered. |
| 10-19 years of age (girls and boys) | Weekly, 1 Iron and Folic Acid tablet  Each tablet containing 60 mg elemental iron + 500 mcg Folic Acid, sugar-coated, blue colour | No corresponding WHO recommendation for women 10-19 years ( Recommendation for adult women and adolescent girls mentioned below) |
| WRA (non pregnant non lactating 20-49 years) | Weekly, 1 Iron and Folic Acid tablet  Each tablet containing 60 mg elemental Iron + 500 mcg Folic Acid, sugar-coated, red colour  All women in the reproductive age group in the pre-conception period and upto the first trimester of the pregnancy are advised to have 400 mcg of Folic Acid tablets, daily | For adult women and adolescent girls daily iron supplementation (30-60 mg) for three consecutive months in a year. Where the prevalence of anaemia in menstruating adult women and adolescent girls is 40% or higher (10)  Weekly Iron 60 mg and Folic Acid 2.8 mg (2800mcg)3 months of supplementation followed by 3 months of no supplementation after which the provision of supplements should restart. If feasible, intermittent supplements could be given throughout the school or calendar year for all menstruating adolescent girls and adult women where the prevalence of anaemia among nonpregnant women of reproductive age is 20% or higher (10) |
| Pregnant women and lactating mothers (0-6 months child) | Daily, 1 Iron and Folic Acid tablet starting from the fourth month of pregnancy (that is from the second trimester), continued throughout pregnancy (minimum 180 days during pregnancy) and to be continued for 180 days, post-partum  Each tablet containing 60 mg elemental Iron + 500 mcg Folic Acid, sugar-coated, red colour | 30-60 mg of iron, with the higher dose preferred in settings where anaemia in pregnant women is a severe public health problem (≥40%), along with 400 µg of folic acid. Daily supplementation throughout pregnancy, beginning as early as possible after conception, is recommended in all settings.(11)  Intermittent use of iron and folic acid supplements by non-anaemic women is a recommended alternative to prevent anaemia and improve gestational outcomes in areas where the prevalence of anaemia among pregnant women is lower than 20%. The suggested dose is 120 mg elemental iron and 2800 µg (2.8 mg) folic acid provided weekly throughout the pregnancy, beginning as early as possible after conception.(12) |

### Supplementary Table 2. The adjusted search terms as per searched electronic databases

| Database | No | Search Query | Results |
| --- | --- | --- | --- |
| PubMed for Q1 19/12/2024 | | | |
|  | #1 | **#1: Intervention Terms** ("iron supplementation" OR "iron intake" OR "iron therapy" OR "iron prophylaxis" OR "iron fortification" OR "iron folic acid supplementation" OR "IFA supplementation" OR "iron tablets") | 9878 |
|  | #2 | **#2: Outcome Terms** ("anemia" OR "iron deficiency anemia" OR "iron deficiency" OR "hemoglobin levels" OR "ferritin levels" OR "hematological outcomes" OR "microcytic anemia" OR "growth" OR "cognitive function" OR "non-hematological outcomes") | 2,686,925 |
|  | #3 | **#3: Population Terms**  ("children" OR "preschool children" OR "school-aged children" OR "6–59 months" OR "5–9 years" OR "adolescents" OR "10–19 years" OR "teenagers" OR "women of reproductive age" OR "15–49 years" OR "pregnant women" OR "pregnancy" OR "antepartum" OR "gestation" OR "maternal") | 3,168189 |
|  | #4 | **#4: Systematic Review Filter**  (“stematic review”; OR “meta-analysis”; OR “Cochrane review”; OR “evidence synthesis”) | 52247 |
|  | #5 | #1 AND #2 AND # 3 AND #4  (("iron supplementation" OR "iron intake" OR "iron therapy" OR "iron prophylaxis" OR "iron fortification" OR "iron folic acid supplementation" OR "IFA supplementation" OR "iron tablets") AND ("anemia" OR "iron deficiency anemia" OR "iron deficiency" OR "hemoglobin levels" OR "ferritin levels" OR "hematological outcomes" OR "microcytic anemia" OR "macrocytic anemia" OR "growth" OR "cognitive function" OR "non-hematological outcomes") AND ("children" OR "preschool children" OR "school-aged children" OR "6–59 months" OR "5–9 years" OR "adolescents" OR "10–19 years" OR "teenagers" OR "women of reproductive age" OR "15–49 years" OR "pregnant women" OR "pregnancy" OR "antenatal") Filters: Meta-Analysis, Systematic Review | 198 |
| Cochrane for Q1 19/12/2024 | | | |
|  | #1 | **#1: Intervention Terms**  ("iron supplementation" OR "iron intake" OR "iron therapy" OR "iron prophylaxis" OR "iron fortification" OR "iron folic acid supplementation" OR "IFA supplementation" OR "iron tablets"):ti,ab,kw | 3469 |
|  | #2 | **#2: Outcome Terms** ("anemia" OR "iron deficiency anemia" OR "iron deficiency" OR "hemoglobin levels" OR "ferritin levels" OR "haematological outcomes" OR "microcytic anemia" OR "growth" OR "cognitive function" OR "non haematological outcomes"):ti,ab,kw | 107161 |
|  | #3 | **#3: Population Terms**  AND ("children" OR "preschool children" OR "school-aged children" OR "6–59 months" OR "5–9 years" OR "adolescents" OR "10–19 years" OR "teenagers" OR "women of reproductive age" OR "15–49 years" OR "pregnant women" OR "pregnancy" OR "antepartum" OR "gestation" OR "maternal"):ti,ab,kw | 312326 |
|  | #4 | #1 AND #2 AND # 3 AND Cochrane Reviews | 102 |
| PubMed for Q2 on 19/12/2024 | | | |
|  | #1 | #1: Intervention Terms (Daily vs. Intermittent)  ("daily iron supplementation"[Title/Abstract] OR "daily iron-folic acid supplementation"[Title/Abstract] OR "daily IFA supplementation"[Title/Abstract] OR "daily iron therapy"[Title/Abstract] OR "iron tablets"[Title/Abstract] OR "intermittent iron supplementation"[Title/Abstract] OR "iron-folic acid supplementation"[Title/Abstract] OR "weekly iron supplementation"[Title/Abstract] OR "iron dosing schedule"[Title/Abstract] OR "iron supplementation regimen"[Title/Abstract] OR "oral iron supplementation"[Title/Abstract] OR "iron and folic acid"[Title/Abstract]) Filters: Meta-Analysis, Systematic Review | 198 |
|  | #2 | **#2: Outcome Terms** ("anemia"[Title/Abstract] OR "iron deficiency anemia"[Title/Abstract] OR "iron deficiency"[Title/Abstract] OR "hemoglobin"[Title/Abstract] OR "ferritin"[Title/Abstract] OR "hematological outcomes"[Title/Abstract] OR "microcytic anemia"[Title/Abstract] OR "growth"[Title/Abstract] OR "cognitive function"[Title/Abstract] OR "non-hematological outcomes"[Title/Abstract]) Filters: Meta-Analysis, Systematic Review | 18808 |
|  | #3 | #3: Population Terms  ("children" OR "preschool children" OR "school-aged children" OR "6–59 months" OR "5–9 years" OR "adolescents" OR "10–19 years" OR "teenagers" OR "women of reproductive age" OR "15–49 years" OR "pregnant women" OR "pregnancy" OR "antepartum" OR "gestation" OR "maternal") Filters: Meta-Analysis, Systematic Review | 664477 |
|  | #4 | #4: Systematic Review Filter  (“stematic review”; OR “meta-analysis”; OR “Cochrane review”; OR “evidence synthesis”) | 52247 |
|  | #5 | **#**1 AND #2 AND # 3 AND #4 | 57 |
| Cochrane for Q2 19/12/2024 | | | |
|  | #1 | ("anemia" OR "iron deficiency anemia" OR "iron deficiency" OR "hemoglobin levels" OR "ferritin levels" OR "haematological outcomes" OR "microcytic anemia" OR "growth" OR "cognitive function" OR "non haematological outcomes"):ti,ab,kw | 107161 |
|  | #2 | ("daily iron supplementation" OR "daily iron-folic acid supplementation" OR "daily IFA supplementation" OR "daily iron therapy" OR "daily IFA tablets" OR "intermittent iron supplementation" OR "intermittent iron-folic acid supplementation" OR "intermittent IFA supplementation" OR "weekly iron supplementation" OR "weekly IFA supplementation" OR "biweekly iron supplementation" OR "biweekly IFA supplementation" OR "iron and folic acid tablets" OR "iron-folic acid prophylaxis" OR "oral iron supplementation" OR "iron supplementation regimen" OR "iron dosing schedule") ti,ab,kw | 3 |
|  | #3 | ("children" OR "preschool children" OR "school-aged children" OR "6–59 months" OR "5–9 years" OR "adolescents" OR "10–19 years" OR "teenagers" OR "women of reproductive age" OR "15–49 years" OR "pregnant women" OR "pregnancy" OR "antepartum" OR "gestation" OR "maternal"):ti,ab,kw | 312326 |
|  | #4 | #1 AND #2 AND #3 AND Cochrane Reviews | 3 |
| PubMed for Q3 on 19/12/2024 | | | |
|  | #1 | ("iron supplementation" OR "iron intake" OR "iron therapy" OR "iron prophylaxis" OR "iron fortification" OR "iron folic acid supplementation" OR "IFA supplementation" OR "iron tablets") Filters: Meta-Analysis, Systematic Review | 365 |
|  | #2 | ("adverse effects" OR "side effects" OR "adverse outcomes" OR "negative outcomes" OR "harmful effects" OR "toxicity" OR "safety concerns" OR "risks" or “complication”) Filters: Meta-Analysis, Systematic Review | 110691 |
|  | #3 | "iron-replete" OR "non-anemic" OR "iron sufficient" OR "iron-sufficient individuals" OR "children" OR "adolescents" OR "women of reproductive age" OR "pregnant women" OR "pregnancy" OR "maternal" OR "antepartum") Filters: Meta-Analysis, Systematic Review | 182551 |
|  | #4 | **#4: Systematic Review Filter** ("systematic review" OR "meta-analysis" OR "Cochrane review" OR "evidence synthesis") | 52247 |
|  | #5 | #1 AND #2 AND #3 AND #4 | 125 |
| Cochrane for Q3 19/12/2024 | | | |
|  |  | ("iron supplementation" OR "iron intake" OR "iron therapy" OR "iron prophylaxis" OR "iron fortification" OR "iron folic acid supplementation" OR "IFA supplementation" OR "iron tablets"):ti,ab,kw | 3469 |
|  |  | ("adverse effects" OR "side effects" OR "adverse outcomes" OR "negative outcomes" OR "harmful effects" OR "toxicity" OR "safety concerns" OR "risks" or “complication”) ti,ab,kw | 417116 |
|  |  | ("iron-replete" OR "non-anemic" OR "iron sufficient" OR "iron-sufficient individuals" OR "children" OR "adolescents" OR "women of reproductive age" OR "pregnant women" OR "pregnancy" OR "maternal" OR "antepartum") ti,ab,kw | 309505 |
|  |  | #1 AND #2 AND #3 AND Cochrane Reviews | 108 |
| Total |  | | **593** |

### Table no 3: AMSTAR-I ratings for each systematic review

| **Review title** | **1.*** | **2.*** | **3.*** | **4.*** | **5.*** | **6.*** | **7.*** | **8.*** | **9.*** | **10.*** | **11.*** | **Total score (out of a maximum of 11)** | **Quality of reviews** |
| --- | --- | --- | --- | --- | --- | --- | --- | --- | --- | --- | --- | --- | --- |
| Effect of daily iron supplementation on health in children aged 4–23 months (Pasricha et al.,) (70) | Yes | Yes | Yes | Yes | No | Yes | Yes | No | Yes | Yes | Yes | 9 | High |
| Micronutrient Supplementation and Fortification Interventions on Health and Development Outcomes among Children Under-Five in Low- and Middle-Income Countries (Tam et al.,) (73) | Yes | Yes | Yes | Yes | Yes | Yes | Yes | No | Yes | Yes | Yes | 10 | High |
| Oral iron supplementation and anaemia in children according to schedule, duration, dose and cosupplementation (Anderson et al., ) (64) | Yes | Yes | Yes | No | No | yes | Yes | No | Yes | Yes | Yes | 8 | High |
| Effect of iron supplementation on physical growth in children: Systematic review of randomised controlled trials (Sachdev et al.,) (72) | No | No | Yes | Yes | No | Yes | Yes | No | Yes | Yes | Yes | 7 | Moderate |
| Effects of daily iron supplementation in primary-school–aged children: systematic review and meta-analysis of randomized controlled trials. (Low et al,.)(69) | Yes | Yes | Yes | Yes | No | Yes | Yes | No | Yes | No | Yes | 8 | High |
| Effects of iron supplementation on cognitive development in school-age children: Systematic review and meta-analysis. (Gutema et al.,)(68) | Yes | Yes | Yes | No | No | Yes | Yes | No | Yes | Yes | No | 7 | Moderate |
| Effect of Oral Iron Supplementation on Cognitive Function among Children and Adolescents in Low- and Middle-Income Countries: A Systematic Review and Meta-Analysis (Chen Z et al.,)(65) | Yes | Yes | Yes | No | No | Yes | Yes | No | Yes | Yes | Yes | 8 | High |
| Intermittent oral iron supplementation during pregnancy (Peña-Rosas JP et al.,) (86) | Yes | Yes | Yes | Yes | Yes | Yes | Yes | No | Yes | Yes | No | 9 | High |
| Daily oral iron supplementation during pregnancy (Peña-Rosas JP et al.,)(77) | Yes | Yes | Yes | Yes | Yes | Yes | Yes | Yes | Yes | Yes | No | 10 | High |
| Daily iron supplementation for improving anaemia, iron status and health in menstruating women (Low MSY et al.,) (74) | Yes | Yes | Yes | Yes | Yes | Yes | Yes | Yes | Yes | Yes | No | 10 | High |
| The benefits and harms of oral iron supplementation in non-anaemic pregnant women: A systematic review and meta-analysis (Watt et al.,) (79) | Yes | Yes | Yes | Yes | No | Yes | Yes | No | Yes | Yes | Yes | 9 | High |
| Iron supplements in pregnant women with normal iron status:  A systematic review and meta-analysis (Hansen et al.,) (80) | Yes | Yes | Yes | Yes | No | Yes | Yes | No | Yes | Yes | No | 8 | High |
| The Effect of Low Dose Iron and Zinc Intake on Child Micronutrient Status and Development during the First 1000 Days of Life: A Systematic Review and Meta-Analysis (Petry et al.,) (71) | No | No | Yes | No | No | No | Yes | No | Yes | Yes | Yes | 5 | Moderate |
| Intermittent iron supplementation for reducing anaemia and its associated impairments in adolescent and adult menstruating women (Gaxiola et al.,) (75) | Yes | Yes | Yes | Yes | Yes | Yes | Yes | Yes | Yes | Yes | No | 10 | High |
| Effects of daily oral iron supplementation during pregnancy (Finkelstein et al.,) (78) | Yes | Yes | Yes | Yes | Yes | Yes | Yes | Yes | Yes | Yes | No | 10 | High |
| Effect of Iron Supplementation on Haemoglobin Response in Children: Systematic Review of Randomised Controlled Trials (Gera T et al.,) (67) | No | Yes | Yes | Yes | No | Yes | No | No | Yes | Yes | No | 6 | Moderate |
| Efficacy of daily versus intermittent oral iron supplementation for prevention of anaemia among pregnant women: a systematic review and meta-analysis (Banerjee et al.,)(85) | Yes | Yes | Yes | No | No | Yes | Yes | Yes | Yes | Yes | Yes | 9 | High |
| Intermittent iron supplementation for improving nutrition and development in children under 12 years of age (De-Regil et al.,)(66) | Yes | Yes | Yes | Yes | Yes | Yes | Yes | Yes | Yes | Yes | No | 10 | High |
| Effects and safety of preventive oral iron or iron+folic acid supplementation for women during pregnancy (Peña‐Rosa and Viteri ) (81) | Yes | Yes | Yes | Yes | Yes | Yes | Yes | Yes | Yes | Yes | Yes | 11 | High |
| Guts, Germs, and Iron: A Systematic Review on Iron Supplementation, Iron Fortification, and Diarrhea in Children Aged 4-59 Months (Ghanchi et al.,) (87) | Yes | No | Yes | No | Yes | Yes | Yes | No | No | No | Yes | 6 | Moderate |

1. Was an 'a priori' design provided? (Yes: the research question and inclusion criteria were established before conducting the review.)

2. Was there duplicate study selection and data extraction? (Yes:.

Was a comprehensive literature search at least two people working independently extracted the data and the method was reported for reaching consensus if disagreements arose.) performed? (Yes: at least two electronic sources were searched; details of the databases, years searched and search strategy were provided; the search was supplemented by searching of reference lists of included studies, and specialised registers, and by contacting experts.)

4. Was the status of publication (i.e. grey literature) used as an inclusion criterion? (Yes: authors searched for reports irrespective of publication type. They did not exclude reports

based on publication from the systematic review. No: the authors stated that they excluded studies from the review based on publication status.)

5. Was a list of studies (included and excluded provided)? (Yes: a list was provided.)

6. Were the characteristics of the included studies provided? (Yes: data on participants, interventions and outcomes were provided, and the range of relevant characteristics reported.)

7. Was the scientific quality of the included studies assessed and reported? (Yes: predetermined methods of assessing quality were reported.)

8. Was the scientific quality of the included studies used appropriately in formulating conclusions? (Yes: the quality, and limitations, of included studies were used in the analysis,

conclusions and recommendations of the review.)

9. Were the methods used to combine the findings of studies appropriate? (Yes: if results were pooled statistically, heterogeneity was assessed and used to inform the decision of

the statistical model to be used. If heterogeneity was present, the appropriateness of combining studies was considered by review authors.)

10.Was the likelihood of publication bias assessed? (Yes: publication bias was explicitly considered and assessed.)

11.Was the conflict of interest stated? (Yes: source of funding or support for the systematic review AND for each of the included studies was clearly acknowledged)

### TABLE 4: EFFECTIVENESS OF IRON SUPPLEMENTATION IN VARIOUS AGE GROUPS

| **Authors and Publication year** | **Origin and number of studies** | **Population** | **Interventions** | **Control** | **Doses / duration**  **/Adherence** | **Study Result (Effect size RR (95%CI)/MD(95%CI), p value n= (number of studies, number of participants)** | **Quality assessment tool** |
| --- | --- | --- | --- | --- | --- | --- | --- |
| Pasricha et al., 2013 (70) | Global  33 studies | Children 4–23 months of age | Daily oral iron supplements | Placebo or no intervention. | **Adherence** not reported (n=11studies), no difference between two groups (n=11) , all doses under supervision (n=6), poor adherence in iron goup (n=2)  **Dose, duration**: Varying doses and duration. Sub group analysis done on doses and duration | **Haematology**  (i)Anaemia: RR 0·61 (0·50,0·74);p <0.001; n= 17, 4825,  (*Anemic = RR 0.60 [0.37, 0.98], n=2, 720; Non-Anemic = RR 1.03 [0.37, 2.91], n=2, 181; Mixed/Unreported = RR 0.60 [0.48, 0.74], n=13, 3924*.)  (ii)Hb= MD 7.22 g/L (4.87, 9.57);**p<0.00001, n=** 26, 5479  (*Anemic=14.14g/L [7.36, 20.92],p<0.0001, n=3, 635; non anemic =MD 11.64 [-5.00, 28.28],p=0.17, n=4,228);Mixed=MD 5.81g/L [3.96, 7.66], p<0.0000)*  (iii) ferritin : MD 21.42 ng/mL) (17.25, 25.58);P<0.00001; n= 23, 4236,  (Anemic = MD 22.24 µg/L [-12.43, 56.91], n=2, 136; Non-Anemic = MD 15.71 µg/L [-0.80, 32.22], n=5, 384; Mixed/Unreported = MD 22.95 µg/L, n=17, 3716)  (ii)ID: RR 0·30 (0·15,0·60); p=0.0006, n=9, 2464  (*Anemic = Not reported; Non-Anemic = RR 0.43 [0.06, 3.28], n=1, 16; Mixed/Unreported = RR 0.30 [0.15, 0.60], n=8, 2448.)*  (iii) IDA: RR 0·14 (0·10,0·22)p<0.00001; n=6, 2145  **Growth**  no differences between children randomised to iron or control in final length (MD –0·13 cm (–0·33 to 0·07) p=0.20, n=7,2470),  length-for-age Z score (MD 0·01 (–0·04 to 0·06) p=0.71, n=8,3237)  final weight (MD –0·02 kg(–0·09 to 0·05); p=0.56; n=8,2702), weight-for-age Z score (MD –0·02 (–0·08 to 0·03); p=0.43; n=8,3237)  weight-for-length Z score (MD 0·03 (–0·06 to 0·12); p=0.50; n=5,2763)  **Development**  **Bayley’s mental development index (score**)- MD 1•65 (–0•63 to 3•94) 0.16 n=6,1093  **(***Anemic = MD 4.46 [-9.32, 18.24], n=3, 113; Non-Anemic = MD 1.49 [-1.08, 4.07], n=5, 325; Mixed/Unreported = MD 0.49 [-2.45, 3.43], n=1, 655.)*  **Bayley’s psychomotor development index (score)**  MD 1•05 (–1•36 to 3•46) 0•39 n=6,1086 | Cochrane instrument  9 studies were judged to be at low risk of bias |
| Tam et al., 2020 (73) | LMICs  197 studies | Under-5 yr children | Vitamin A/ Zinc/ Iron/ IFA Supplementation  *(Only Iron/IFA supplementation verus placebo included in the table)* | placebo/ no supplementation/  standard of care | No information on adherence.  **Doses: No subgroup analysis on different doses**  Duration-- <3 months, 3–5 months,  and 6–12 months | **Haematological**  **(a) Iron Supplementation**  (i)Anaemia: RR 0.55, (0.44,0.70); p < 0.00001, n= 14,1511  (ii)Hb: MD 6.02 g/L, (4.28,7.76),p< 0.00001,  (iii)plasma/serum ferritin: MD 20.48 µg/L, (13.41,27.55), p < 0.00001)  (iv)ID: RR 0.21, (0.12,0.39), p <0.00001)  (v) IDA: RR 0.14, (0.04, 0.54),p = 0.004)  **Anemia risk:**  (vi) greater reduction1–5-monthRR 0.41; (0.28,0.60) p<0.00001, n=7,1576, and 6–11-month age groups (RR 0.59; (0.47,0.75), p<0.0001,n= 3,548) compared to 24–59-month RR 0.67; (0.39,1.15), p=0.14;n=1,194)p for subgroup differences = 0.22  (vi) Greater reduction in non -anaemic at baseline RR 0.46; (0.31,0.68); p<0.00001;n=4,806) than anaemic (RR 0.72;(0.53,0.96), p=0.03; n=3,432), p for subgroup differences = 0.07  **(b) IFA supplementation**  (i)Anaemia RR 0.80, (0.66, 0.97),p=0.02, n=4,614  ii)Hb concentration MD 3.06 g/L, (1.16,4.97), p=0.002; n=4  **Growth;**  **(i) Stunting** : No difference RR 0.96 (0.77,1.18);p=0.67; n=4,1138  **(ii)Wasting:** No difference RR 1.21 (0.82,1.79); p=0.34; n= 3,903  **(iii) Height**: MD -0.06cm (-0.23,0.10), p=0.45, n=13,2326  **(iv) Weight**: MD 0.00(-0.06,0.07), p=0.95, n=13,2864  **(v) Length for age (z-score**): MD -0.01(-0.20,0.18),p=0.89, n=13,2904  **(vi) HAZ:** MD 0.00(-0.06,0.06),p=0.88, n=13,2822  (**vii) WAZ**: MD 0.01(-0.03,0.06),p=0.54, n=13,2836  **Development**  **Mental Development -** SMD 0.14, 95% CI 0.01 to 0.28; p = 0.04, n=4,453)  **motor development** -SMD 0.28, 95% CI 0.15 to 0.40; p < 0.0001, n=3,517), | Cochrane Risk of Bias Tool  The majority of the included studies had a low risk of bias across many domains, indicating a low overall risk of bias. |
| Andersen CT, 2023 (64) | Global  129 trials | children and adolescents aged <20 years | Daily Oral iron Intervention [Frequent (3–7/week) and intermittent (1–2/week) iron regimens] | placebo/No intervention | **Adherence:** No quantitative assessment of adherence done as included studies did not report adherence uniformly.  Subgroup analysis based on different doses was done for each age group | **Overall Increases**  **(i)**Hb: WMD 6.3 g/L (5.5, 7.1);p<0.001; n=167  (  (ii) Ferritin: WMD 18.5 ng/mL(16.1,20.9); p<0.001;n=107;  (iii) AnemiaRR 0.61 (0.55,0.67) p<0.001; n=69  (iv)ID RR 0.30 (0.24,0.37)p<0.001; n=48;  (v)IDA RR 0.20 (0.13,0.31); p<0.001; n=27  **By Age Group:**  **(i)Children 0-5 months:**Hb: WMD 5.2 g/L(3.7, 6.7) n=39; Ferritin: WMD 25.1 ng/mL(19.7, 30.5) n=33;; Anemia: 43% (RR 0.57, (0.48, 0.70) n=18; ID: 77% (RR 0.23 ((0.15, 0.35)n=19; IDA: 83% (RR=0.17, (0.10, 0.31) n=17)  **(ii)Children 6-23 months:**Hb: WMD 5.1 g/L(3.3,6.9); n=31; Ferritin: WMD14.4 ng/mL(8.7, 20.0);n=33 Anemia: 23% (RR 0.77, (0.67, 0.90)n=23); ID: 62% (RR 0.38(0.27, 0.53)n=15; IDA: 68% (RR0.32 (0.14, 0.69) n=5)  **(iii)****Children 2 to <5 years:**Hb: WMD 5.7 g/L(3.8, 7.7);n=25 Ferritin: WMD19.5 ng/mL(11.3, 27.7);N=10, Anemia: -24% (RR0.76, (0.53, 1.09)n=6); ID: -69% (RR0.31 (0.14, 0.67) n=4), IDA RR 0.19 (0.06, 0.56)n=4  **(iv) Children ≥12 years:** Hb: WMD 6.6 g/L(4.4,8.7);n=21 Ferritin: WMD 10.9 ng/mL(9.1,12.7);N=20, Anemia: - (RR 0.49, (0.31, 0.79)n=5); ID: RR0.30 (0.11,0.80) n=3), IDA RR 0.09 (0.01, 1.64)n=1  **By Baseline Anemia Status:**  **(i)Anemic:**Hb: WMD10.6 g/L (7.3, 13.9), n=33; Ferritin: WMD 13.1 ng/mL (8.0, 18.2); n=17; Anemia: 65% (RR 0.35, (0.26, 0.47) n=9); ID: 81% (RR 0.19, (0.06, 0.61) n=5 ; IDA: 85% (RR 0.15 (0.02, 1.26), n=1)  **(ii)Mixed Anemic and Non-anemic**:Hb: WMD 6.5 g/L (5.4, 7.5), n=54; Ferritin: WMD 22.7 ng/mL (17.4, 27.9), n=30; Anemia: -37% (RR0.63 (0.55, 0.71) n=40) ; ID: -74% (RR0.26 (0.19, 0.36) n=23); IDA: -87% (RR0.13 (0.08, 0.22)n=13)  **(iii)Non-anemic:**Hb: WMD 5.1 g/L (3.5, 6.7),n=28; Ferritin: WMD 18.7 ng/mL (13.5, 23.9),n=20;Anemia: 0.62 (0.41,0.93)); n=10; ID: 0.20 (0.11,0.36); n=5; IDA: 0.21(0.06,0.70)); n=4  **Regions &Genders**:  Ages<5 months to >12 years, both genders; future anemia risk – 37%, and ID and IDA – 80%.  **Frequent (3-7 times/week)**  (i) Hb: WMD 6.6 (5.6,7.6); n=132  (ii)Ferritin (ng/mL): WMD 20.9; (18.2,23.7); n=88  (iii) Anaemia; RR 0.62; (0.56,0.69); n=57  (iv)ID; RR 0.31; (0.24, 0.38); n=44  (v)IDA; RR 0.20; (0.13,0.32); n=24  **Intermittent (1–2 times/week)**  (i)Hb: WMD 4.8 (3.4,6.2); n=32  (ii)Ferritin (ng/mL): WMD 6.7; (3.7,9.6); n=18  (iii)Anaemia; RR 0.61; (0.41,0.92); n=10  (iv)ID; RR 0.24; (0.09,0.66); n=4  (v) IDA; RR 0.22; (0.06,0.77); n=3  **Growth**:  (i) HAZ score: Anaemic– increased RR 0.20 (0.01, 0.40) 2 studies;  non-anemic RR -0.02 ;(-0.18, 0.14); N=3) or mixed groups (RR 0.00 (-0.03, 0.02); N=25- no effect | Cochrane Risk of Bias Tool  48/ 129 trials (37%) high risk of bias for at least one of the criteria.  24/129 trials (19%) high risk of bias for inadequate outcome data. |
| Sachdev H, Gera T, Nestel P, 2006 (72) | Global 25 studies  (13 from Asia) | infants and toddlers and older Children | oral or parenteral iron supplementation, or iron-fortified formula milk or cereals. | Placebo, except in trials where iron was given parenterally, in which case the control group did not receive a placebo. | **Adherence**: Not mentioned  **Duration:**03 months (n=8), >3-< 6months ( n=10), >6 months (n=8)  **Doses: No subgroup analysis on different doses** | **Growth**  i) Weight-for-age: SMD 0.13 (-0.05, 0.32); P=0.14, n=22,4327  ii) Weight-for-height: SMD 0.27 (-0.14, 0.69); P = 0.18; n=8, 1246  iii) Height-for-age: SMD 0.06(-0.07, 0.20); P = 0.38; n= 21,3935  iv) Mid upper-arm circumference: SMD 0.0 (-0.20, 0.20); p=0.991;n=12, 1163  v)TRiceps Skinfold thickness, MUAC, Head circumference & sub scapular no Difference | No details |
| Low et al., 2013 (69) | low- or middle-income  32 studies | children aged 5–12 years | daily iron supplementation (≥ 5 d/wk) | No supplementation/Placebo/Co supplementation | **Adherence:** >90% (n=4), >80% (n=7), 72% (n=1), 50% (n=1). Six studies excluded participants with <66%-90% adherence. Five studies reported similar adherence in iron and control groups. Metanalysis not possible due to inconsistent reporting  **Dose and Duration:** Subgroup analysis | **Hematological:**   (i) Anemia: 50% (RR 0.50; (0.39,0.64) p<0.001, n=7, 1763 (ii) ID: 79% (RR 0.21 (0.07,0.63) P=0.006, n=4, 1020. (iii) Ferritin MD28.45 μg/L (18.03,38.86) p< 0.001, n=14, 3612 (iv) Hb: MD 8.38 g/L (6.21 to 10.56) p< 0.001; n=28,6545   **Development:**   (i) Global Cognitive Scores: Higher in supplemented children SMD 0.50, (0.11, 0.90), p = 0.01, n = 9, 2355) (ii) IQ Scores: No significant effect (MD 5.47, (−3.24,14.18), p = 0.2,n = 5, 1874) but anemic at baseline had significant improvements-(MD 4.55, 95% CI 0.16 to 8.94; p = 0.04, n= 3,186) (iii) Maze Test Performance: Improved (MD 1.30, (0.9,1.7), p < 0.001, n = 4, 288) (iv) Clerical Task Scores: Higher (SMD 0.44, (0.14,0.75), p = 0.005,n = 4,290)   **Growth:- No benefit of iron supplemention on height , weight associated Z scores**  (i) (Height = MD -0.37 [-1.14, 0.40], p=0.3, n=5, 1111  (iiHAZ = MD 0.09 [0.01, 0.17], p=0.03, n=5, 1318  (iii) Weight = MD 0.12 [-0.50, 0.75], p=0.7, n=5, 1111  (iv) WAZ = MD 0.10 [-0.03, 0.23], p=0.1, n=5, 1318   (i) HAZ: MD 0.09, (0.01,0.17),p=0.03, n=5, 1318 | RoB  There are 4 studies with low risk of bias |
| Gutema et al., 2023 (68) | Global  13 studies | school-age children (age ranging between 6 to 12 years) | In most of the studies supplements were provided at least 5 days / week. Children were supplemented once or twice weekly, four times per week or daily in three studies. Five included studies supplemented children for 2–3 months. Five studies provided the supplement for 4 months and three studies for 8 months to a year. | Placebo or no intervention | **Adherence:** Reported in 04 studies, all had adherence more than 70%.No difference between groups  **Dose and Duration**: Ranged from 2-60 mg elemental iron  Subgroup analysis based on duration of supplementation | **Hematological**  i) Hb: SMD 1.08 (0.68 to 1.49), **p < 0.001;** n = 11, 3613)  ii) Serum Ferritin: SMD 1.93 (-0.27, 4.14), p = 0.08; n = 4, 2570)  iii) Transferrin: SMD 1.01 (-0.50,2.51), p = 0.19; n = 4, 1970)  **Cognitive Outcomes:**  i) Intelligence: SMD 0.46 (0.19,0.73), **p < 0.001**; n = 10, 3105)  ii) Attention and Concentration: SMD 0.45 (0.09,0.80)**, p = 0.01**,n =3, 339)  iii) Memory: SMD 0.45 (0.04, 0.69), p < 0.001, n =5, 718)  **Subgroup Analysis (Anemic Children):**  i) Intelligence: SMD 0.79 (0.41,1.16), **p = 0.001**, ; n = 3,236)  ii) Memory: SMD 0.47 (0.13,0.81), **p = 0.006**; n = 3, 145)  iii) School Achievement: SMD 0.00 (-0.21,0.21), p = 1.00; n =5, 2272) | Cochrane risk of bias assessment tool (RoB2)  Six studies were considered as high RoB.  Four represent some concern.  Three have low RoB |
| Chen Z et al., 2022 (65) | LMICs  9 studies | Children and adolescents (6-19 yrs) | Iron supplementation | Placebo or no intervention | **Adherence:** Not reported  **Dose**  Doses ranges from <60-≥60 mg/day  **Duration :** The median intervention duration was 4.9 months (varied from 3-8.5 months) | (i) Short term memory: SMD 0.74 ( -0.15, 1.63), p=0.104;n=4 (ii) Long term memory: SMD = 0.94, (−0.90, 2.77), p = 0.315; n=2 (iii) School performance = SMD 0.42 (-.33,1.18) P=0.275; n=4 (iv) Intelligence test scores: SMD 0.47;( 0.10,0.83); P=0.012, n=8,   (a)Age> 11 years :SMD 1.18,( 0.33, 2.04), p = 0.007  (b)daily supplementation dose ≥60 mg/day: SMD 0.91,( 0.38, 1.45), p = 0.001,  I≥4 months :SMD 0.81,( 0.38, 1.25), p < 0.001  (d) anemic:SMD 1.01, (0.34, 1.68), p = 0.003,  non-anemia group: SMD = 0.68, (0.17, 1.18), p = 0.009, | Cochrane risk of bias assessment tool (RoB2)  5 studies low risk  2 unclear risk  2 high risk |
| Gera et al., 2007 (67) | Global 55 trials (23 from Asia) | Children 0-18 year | Iron supplementation | Control/ PLacebo | **Not reported** | **Hematological**   (i) Hb: WMD 0.74 g/dL (0.61,0.87) p<0.001, (ii) <11 g/dL Hb: WMD 1.10 (0.79, 1.41)p<0.001, (iii) >11 g/dL Hb: WMD 0.49 (0.39, 0.59) p<0.001, (iv) <24 months: WMD 0.56 (0.36, 0.76) p<0.001, (v) <60 months: WMD 0.59 (0.43, 0.75) p<0.001, (vi) >60 months: WMD 0.88 (0.67, 1.08) p<0.001, (vii) Developing countries 0.78 (0.64, 0.93) <0.001, |  |
| De‐Regil et al., 2011 (66) | Developmental Countries  33 trials | children under the age of 12 | Intermittent iron supplementation, alone or in combination with other vitamins and minerals, | Placebo, no intervention, or daily intervention. | **Adherence:**  2 studies reported children of intermittent group shows similar level of adherence of intermittent placebo group. (RR 1.04, 95% CI 0.98 to 1.09)  **Doses**:  Intermittent iron dose per week (25 mg or less; greater than 25 mg to 75 mg; greater than 75 mg)  **Duration:**  0-3 months and >3 months | **Intermittent group:**  (i)Higher anaemia RR 0.51, (0.37–0.72)n=10,1824  (ii)Higher ID RR 0.24, (0.06,0.91); n=3, 431  (iii)Ferritin MD 14.17 µg/L (3.53,24.81), n=5, 550  (iv)Higher haemoglobin MD 5.20 g/L, (2.51,7.88), n=19, 3032 | GRADE  Anemia, Haemoglobin, Ferritin= Low,  ID= Very Low |
| Low MSY et al., 2016 (74) | Global  67 studies | Non‐pregnant women of reproductive age (menstruating women 12 – 50 years) | Daily oral supplementation with any dose for at least 05 days a week with or without co intervention. | Control or Placebo | **Adherence**: Not reported (n=34); other studies reported adherence in heterogenous ways, meta‐analysis not done.  Participants of iron group didn’t have poorer adherence than placebo group.  **Dose :**  Varying doses of elemental iron. **Duration of iron supplementation:**  ≤ 1 month, 1-3 months,> 3 months. | (i)Anemia: RR 0.39 (0.25,0.60),n = 10,3273)  (ii)Hb: MD 5.30 g/L (4.14,6.45), n = 51, 6861)  (iii)ID: RR 0.62 (0.50,0.76), n =7, 1088)  (iv) Feritin MD 10.27 ng/mL (8.90,11.65)p<0.0001, n= 42,3881  (v) Serum Iron SMD0.47, (0.19,0.74); P < 0.00001, n=17, 902  **Growth:**  Height, weight, BMI- no effect | GRADE  Anemia, ID= Moderate,  Hb= High |
| Fernández-Gaxiola, Luz Maria De-Regil., 2019 (75) | Global  25 studies | WRA (women beyond menarche and prior to menopause who were not pregnant or lactating and did not have a known condition that impeded the presence of menstrual periods) | Iron supplementation (one, two or three times a week on non-consecutive days | placebo, no intervention or daily supplementation | **Adherence:**  2 studies examine adherence. No evidence of women in intermittent group adhere to intervention better the no supplement group (RR 0.99 (0.96, 1.02); n=2, 417.  **Dose:**  Doses ranging from ≤60 mg/week- ≥60 mg/week  **Duration:**  one, two or three times a week on non-consecutive days | **Intermittent vs No intervention/Placebo**  **Haematological:**  (i)Anemia: RR 0.65; (0.49,0.87); p<0.001;n=11, 3135  (ii)Hb: MD 5.19 g/L,( 3.07,7.32); p<0.001 ; n=15, 2886)  (iii)Ferritin (MD 7.46 μg/L, (5.02,9.90); p= 0.09 ;n=7,1067  (iv)ID (RR 0.50, (0.24,1.04); p<0.001;n=3,624  (v)IDA (RR 0.07, (0.00,1.16); n=1,97  **Others**  (i)All‐cause morbidity (RR 1.12, (0.82, 1.52); n=1,119 | GRADE  Anemia, ID, IDA, Ferritin, All cause morbidity= Low.  Hb, Any adverse effect= moderate. |
| Peña-Rosas JP et al., 2015 (77) | Global  61 studies | Pregnant Women | daily iron and iron plus folic acid supplementation/ Vitamin Supplementation/Mineral Supplementation | Folic Acid/Placebo/ Vitamin Supplementation/Mineral Supplementation | Not reported | **Hematological (Iron supplementation)**  (i)Anemia at term: RR 0.30; (0.19,0.46);n=14,2199).  (ii)Iron-deficiency anemia: RR 0.33; (0.16, 0.69); n=6, 1088).  (iii)Severe anemia (2^nd^& 3^rd^ trimesters): Reduced risk (RR 0.22; (0.01, 3.20);n=9,2125).  (iv)ID at term: Reduced risk RR 0.43; (0.27, 0.66);n=7, 1256).  **Birth outcomes**  (i)Low birth weight: No significant effect RR 0.84; (0.69,1.03); n=11,17613).  (ii)Birth weight: No significant increase MD 23.75 g; (-3.02,50.51); n=15, 18590).  (iii)Preterm birth: No significant effect RR 0.93; (0.84,1.03);n=13, 19286).  (iv)Neonatal death: No significant effect RR 0.91; (0.71,1.18); n=4, 16603).  (v)Congenital anomalies: No significant effect RR 0.88; (0.58, 1.33);n=4, 14636). | GRADE  LBW, BW, PTB= low  Neonatal death, Maternal anaemia at term, Maternal IDA at term, side effects and Severe anaemia at any time during second and third trimester = very low grade |
| Watt et al., 2024 (79) | Global  (23 studies) | Non anemic Pregnant women | oral iron supplementation | placebo or no supplement | Not reported | **Hematological**  (i)Maternal Anemia: OR 0.33 (95% CI = 0.20 - 0.56, p<001) | ROB & NOS  “Low” only 2 studies, “Some concerns” for 13 studies and “High risk” for 6 studies |
| Hansen et al., 2023 (80) | Global  (08 studies) | non-anemic iron replete pregnant women | - Daily oral iron supplements | No supplementation/ Placebo | **Adherence:**  3 study reported adherence. 2 of them reported more than 60% adherence while 1 study reported very low (6%)  **Dose and Duration:**  Doses ranging from ≤30 mg/day- ≥60 mg/day | (i)IDA at term :RR 0.51, (0.38,0.70), n=4, 1670  (ii)ID at term: RR 0.74, (0.60–0.92), n=4,1663  (iii) Hb>130 g/L between the groups: RR 0.94, (0.09,10.07), n=3, 1346 | GRADE  Maternal ID= Very Low  Hb, SGA, PTB= low  Maternal IDA , LBW= Moderate |
| Petry et al., 2016 (71) | 90 studies | women (pregnant and lactating)  children (6–23 months) | - Daily iron and zinc dose did not exceed 15 mg and 10 mg, respectively for children - 45 mg and 21 mg, respectively for women | Placebo/Non Iron Supplements | **Not reported** | **Hematological**  **Pregnancy** (iron or zinc)-no effect on birth outcomes.  **Children 6-23 months**  (i)Hb–MD 4.1 g/L(2.8,5.3),p<0.001; n=30,6569  (ii) serum ferritin –MD 17.3 µg/L.( (13.5,21.2), p<0.0001; n=21, 4291,  (iii) Anemia-RR 0.59(0.49;0.70) p<0.0001, n=22, 5647),  (iv)ID RR 0.22(0.14;0.35) p<0.0001;n=13, 3698)  (v) IDA RR 0.20(0.11;0.37), p<0.0001; n=8, 3464)  (vi)Reduces risk of anemia-41%, ID-78%, IDA- 80%.  **Growth**  (i)no effect on growth or psychomotor development. | GRADE  Hb = moderate,  anaemia = low, IDA  = high, ID = high, diarrhoea  = not assessed |
| Finkelstein et al., 2024 (78) | Global  57 trial | Pregnant women | Daily Iron supplementation or IFA with cointerventions like education | No Iron/Placebo/no IFA | **Not reported** | **Pregnancy**  (i)Reduce maternal anemia: RR 0.30 [0.20,0.47]; n=14, 13,543  (ii) ID at term: RR 0.51 [0.38,0.68]; n= 8, 2873  (ii)Maternal IDA at term: RR 0.41 [0.26, 0.63]; n=7, 2704  (iv)Maternal mortality: minimal /no change ;RR 0.57 [0.12,2.69]; n=3, 14,060  **Birth Outcome**  (v)Reduced LBW: RR 0.84, (0.72, 0.99); n=12,18,290;  (vi)Increased Birthweight: MD 24.9 g, (-125.81, 175.60); n=16, 18,554,  (vii)Preterm birth rates RR 0.93(0.84, 1.02); n=11 , 18,827;  (viii)neonatal death rates RR 0.98, (0.77,1.24);n= 4, 17,243; | GRADE  **Maternal Outcome**  ID, Adverse effect- Low  Anaemia at term, maternal Malaria- Moderate.  IDA , Maternal Death, , Severe Anemia- Very low  **Infant Outcome**  LBW, Preterm,  Neonatal Death, Congenital Anomalies- Low  Birth weight- Moderate |
| Peña‐Rosa and Viteri 2009 (81) | Global  49 trial | Pregnant women | Daily use of iron or iron+folic acid supplements | Placebo/Usual care/Co-intervention or intermittent supplementation) | **Dose and duration:**  intermittent (weekly or  twice weekly)  **Adherence:**  Not reported | **Daily IFA vs no intervention/placebo:**  (i)Daily IFA group less anaemic at term(8.2% vs 35.5%) RR 0.27; (0.12,0.56);  (ii) ID at term: RR0.24 [0.06, 0.99], n=1,131  (iii)IDA at term: RR 0.43 [0.17, 1.09], n=1.131  **Daily Iron vs no intervention/placebo:**  (i)Hb at term: MD 8.83 g/L [6.55, 11.11], n=17,2463  (v) anaemia at term RR 0.27; (0.17,0.42); n=14,4390  (ii) ID at term: RR0.44 [0.27, 0.70], n=6,1108  (iii)IDA at term: RR 0.33 [0.16, 0.69], n=6.1667 |  |

### Table 5: Daily versus intermittent iron prophylaxis on haemoglobin ferritin levels in various age groups

| **Authors and Publication year** | **Origin** | **Population** | **Intervention** | **Control** | **Doses / duration**  **/Adherence** | **Outcome** | **Quality assessment** |
| --- | --- | --- | --- | --- | --- | --- | --- |
| De‐Regil et al., 2011 (66) | Developmental Countries  33 trials | children under the age of 12 | Intermittent iron supplementation, alone or in combination with other vitamins and minerals,  (9 trials twice/week, 2 trials 3times/week, 22 trials once/week) | Placebo, no intervention, or daily intervention. | **Adherence:**  2 studies reported children of intermittent group shows similar level of adherence of intermittent placebo group. (RR 1.04, 95% CI 0.98 to 1.09)  **Doses**:  Intermittent iron dose per week (25 mg or less; greater than 25 mg to 75 mg; greater than 75 mg)  **Duration:**  0-3 months and >3 months | **Intermittent group:**  (i)Higher anaemia RR 1.23, (1.04,1.47)n=6,980  (ii)Higher ID RR 4.00, (1.23,13.05); n=1, 76  (iii)No difference Ferritin MD -4.19 µg/L, (-9.42,1.05), n=10, 902  (iv)No difference haemoglobin MD –0.60 g/L, (–1.54,0.35), n=19, 2851  **Side effects**  No difference in both the group RR 0.60, (0.19 to 1.87), n=4,895 | GRADE  Anemia, Haemoglobin, Ferritin= Low,  ID= Very Low |
| Fernández-Gaxiola, Luz Maria De-Regil., 2019 (75) | Global  25 studies | WRA (women beyond menarche and prior to menopause who were not pregnant or lactating and did not have a known condition that impeded the presence of menstrual periods) | intermittent iron supplementation  19 studies provided once weekly supplementation | daily iron supplementation. | **Adherence:**  2 studies examine adherence. No evidence of women in intermittent group adhere to intervention better the no supplement group (RR 0.99 (0.96, 1.02); n=2, 417.  **Dose and Duration:**  Doses ranging from ≤60 mg/week- ≥60 mg/week  **Duration:**  one, two or three times a week on non-consecutive days | **Haematological:**  **Intermittent vs Daily**  (I)anaemia: RR 1.09, (0.93,1.29); p=0.34; n= 8, 1749  (ii)Hb levels MD 0.43 g/L,( −1.44, 2.31); p<0.001; n=10,2127;  (iii)Ferritin: MD-6.07 μg/L, (−10.66, −1.48); p=0.01, n=4,988  **iv) ID**:RR 4.30, (0.56 to 33.20); n=1, 198  **Side effects**  Intermittent supplements RR 1.98, (0.31,12.72), n=3,630 were less adverse effects than daily regime (RR 0.41, (0.21,0.82);n= 6, 1166) | GRADE  Anemia, ID, IDA, Ferritin, All cause morbidity= Low.  Hb, Any adverse effect= moderate. |
| Banerjee et al., 2024 (85) | Global  26 studies | Pregnant Women | Daily Iron supplementation. | Intermittent  ( 20 trials- iron once a week) | **Dose and duration:**  The median intermittent iron dose was 120 mg/day and daily iron dose was 60 mg/day.  **Adherence:**  Not reported | **Hematological: (Daily vs intermittent)**  **(i)Hb**SMD 0.51; (-0.23,1.24), p = 0.18; n=25, 4120 (no sig difference);Non anemic: SMD 0.01 (-0.51,0.53), p=0.97;n=18, 2291,;Anemic: SMD: 0.77, (−0.45,1.99). p=0.22;n=9,1233  **(ii)Ferritin levels** higher in daily group: SMD 0.85; (0.15,1.54), p = 0.02; n=16, 2132  **Hematological: (Daily vs weekly)**  Hb: SMD 0.89(0.07,1.84)  **Side effects:**  Daily group had higher odds of side effects:  (i)Nausea: adjusted OR 3.56, (2.23,5.69), p < 0.001; n=10,995  (ii)Diarrhea: Adjusted OR 5.40, (1.90,15.33), p = 0.002, n=10,995  (iii)Constipation: Adjusted OR 1.95, (1.21,3.14), p = 0.006,n=3, 841  (iv) Vomitting: Adjusted OR 3.22 (0.94,10.95), p = 0.06, n=3, 841 | GRADE  Five studies were classified as having a low risk of bias in all domains, while 19 studies were classified as having a high risk of bias for endpoint haemoglobin levels. |
| Peña-Rosas JP et al.,2015 (86) | Global  21 studies | Pregnant women | Intermittent supplementation of Iron/Iron+ folic acid /other vitamins and minerals  Most of the intermittent regimens involved women taking supplements on one day each week (usually two tablets on the same day each week). | Placebo/Usual care/Co-intervention (Daily Supplementation) | **Adherence:**  Not reported  **Dose and duration:**  intermittent (weekly or  twice weekly)  **Weekly dose:** ranging from 100-180mg/week  **Daily dose:** ranging from 40mg-120 mg/day | **Intermittent vs daily Iron**  **Birth outcomes :**  (i)Low Birthweight: RR 0.82, (0.55, 1.22), n =8, 1898).  (ii) Infant Birthweight:MD 5.13 g, (-29.46,39.72), n = 9, 1939).  (iii)Premature Birth: RR 1.03, (0.76, 1.39),n = 5, 1177).  (iv)Neonatal Death: RR 0.49, (0.04, 5.42), n =1, 795).  (v)Congenital Anomalies: Not reported in any studies.  **Maternal Outcomes:**  (i)Anemia at Term: RR 1.22, (0.84,1.80), n = 4,676.  (ii) Hb >130 g/L in second or third trimester (RR 0.53, (0.38,0.74), n = 15, 2616; not a primary outcome).  (iii)Iron-Deficiency Anemia at Term: No significant difference (RR 0.71, (0.08, 6.63), n = 1, 156).  (iv)Hemoconcentration (Hb>130g/L) in 2^nd^ and 3^rd^ trimester RR 0.53; 95% CI 0.38 to 0.74; n =15, 2616  **Side effect:**  (i)Side Effects: Fewer in intermittent supplementation group (RR 0.56, (0.37, 0.84),n =11, 1777). |  |
| Peña‐Rosa and Viteri 2009 (81) | Global  49 trial | Pregnant women | Daily use of iron or iron+folic acid supplements | Placebo/Usual care/Co-intervention or intermittent supplementation) | **Dose and duration:**  intermittent (weekly or  twice weekly)  **Adherence:**  Not reported | **Intermittent vs Daily IFA**  (i)Hb levels (>130g/L) RR 0.54(0.18,1.58).  (ii) Intermittent Group are less likely to have haemoconcentration compared to daily(7.75%versus 19.31%); RR 0.41; (0.21,0.80)  (iii)Anemia: RR 1.20 [0.78, 1.83]  **Side effect:**  Any sde effect: RR 0.69 [0.45, 1.04], n=7,1307  Diarrhoea: RR 0.94 [0.38, 2.34], n=4,553  Constipation: RR 0.99 [0.48, 2.06], n=4,553 Nausea: RR 0.59 [0.30, 1.16], n=5,854  Vomiting: RR 1.44 [0.82, 2.53], n=5,854 |  |

### Table 6: Adverse effects with iron supplementation in various age groups

| **Authors and Publication year** | **Origin and number of studies** | **Population** | **Interventions** | **Control** | **Study Result** | Assessment Tool |
| --- | --- | --- | --- | --- | --- | --- |
| Pasricha et al., 2013 (70) | Global  (33 studies) | Children (4–23 months old) | Daily oral iron supplements | placebo/no supplement | **Clinical :**  (i)Fever RR1 1·08 (0·79 to 1·47); p=0.63; n=2  (ii) vomiting RR1·38 (1·10 to 1·73)p=0.006; n=3,1020,  (ii) ARI RR 1.04, (0.92,1.19); p=0.51, n=2,944;),  (iv) Diarrhoea RR 1.03, (0.86,1.23); p=0.78; n=6,1697;), (v) Constipation RR 0.54, (0.05-5.83); p=0.49; n=2,570),  (vi) Any side effects (RR1.10, (0.98,1.25); p=0.12, n=3, 912;  (vii) Diarrhoea (Prevalence): 0.68 [0.37, 1.27] (Anemic) , 0.66 [0.17, 2.57] (Non-anemic)  **Growth**:  **Iron supplement group**   i. No significant differences in length for age or weight for age; ii. Reduced length gain SMD –0.83, (–1.53, –0.12); p=0.02; n=8, 868) iii. weight gain SMD–1.12, (–1.91, –0.33); p=0.0005; n=8,868 iv. Stunting (RR1.10,( 0.92,1.32); p=0.29; n=3, 1504;), v. Wasting (RR 1.03, (0.65,1.64); p=0.89; n=3,1504;) vi. >3 months iron supplementation: SMD -2.39; 95% CI: -4.37, -0.41; p=0.02 | Cochrane Instrument |
| Sachdev H, Gera T, Nestel P. 2006 (72) | Global 40 studies  (13 from Asia) | Infants and toddlers | oral or parenteral iron supplementation, or iron-fortified formula milk or cereals. | Placebo, except in trials where iron was given parenterally, in which case the control group did not receive a placebo. | **Growth**:Slight negative effect on linear growth from developed countries SMD = -0.27; (-0.49, -0.05); P = 0.018  iron supplementation>6 months duration SMD = -0.13; (-0.24, -0.01); P = 0.039. | No details |
| Ghanchi et al., 2019 **(SR) (87)** | Global  (19 studies) | Children aged 4–59 month | oral iron supplementation or iron fortification | any placebo or control group | **Iron-supplemented and control groups.**  **Diarrhoea**  7/19 (37%) studies found a significant increase  12/19 trials (63%) found no difference  **iron-replete group:** 2 studies found an increase in bloody diarrhoea. | Cochrane risk of bias tool   - 9/19 (47%) “high” quality - 8/19 (42%)“adequate,” - 2/19 (11%) “low” quality. |
| Andersen CT, 2023 (64) | Global  129 trials | children and adolescents aged <20 years | Intervention =Frequent (3–7/week) and intermittent (1–2/week) iron regimens | placebo/control | **Clinical*-***no significant effect on infections, although small effects on diarrhoea or malaria could not be excluded. |  |
| Low et al., 2013 (69) | low- or middle-income  32 studies | children aged 5–12 years | daily iron supplementation | No supplementation/placebo/co-supplementation | **Clinical**  (i)malaria parasitaemia :No difference RR 1.10, (0.94, 1.29); p = 0.9, n =4, 919)  (ii)No difference in GI adverse effects (Diarrhoea, Constipation &Vomitting) | RoB  There are 4 studies with low risk of bias |
| Fernández-Gaxiola, Luz Maria De-Regil., 2019 (75) | Global  25 studies | WRA | Iron supplementation (one, two or threetimes a week on non-consecutive daysverus daily supplementation | No intervention | Harmful side effects: Less in intermittent group: RR 0.41,I (0.21 to 0.82);n= 6, 1166 | GRADE |
| Low MSY et al., 2016 (69) | Global  67 studies | Non‐pregnant women of reproductive age (menstruating womenaged 12 to 50 years) | Daily oral supplementation with any dose for at least 05 days a week with or without co intervention. | Placebo | **(i)Overall Side Effects**: No significant increase(RR 2.14, (0.94, 4.86), P < 0.00001; n = 7, 901).  **(ii)****GI side effects**: Increased prevalence in iron group RR 1.99, (1.26,3.12), p=0.12; n = 5, 521).  **(iii) Loose Stools/Diarrhea:** Increased prevalence RR 2.13, (1.10,4.11),p=0.31; n = 6,604).  **(iv)Hard Stools/Constipation:** Increased prevalence (RR 2.07, (1.35,3.17), p=0.77; n = 8, 1036).  **(v)Abdominal Pain:** Increased prevalence (RR 1.55, (0.99,2.41), n = 7, 1190).  **(vi)Nausea:** No significant increase RR 1.19, (0.78 to 1.82), p=0.51; n = 8, 1214). | GRADE  Over all side effect, GI, Abdominal Pain= Low.  Loose Stools/Diarrhea, Hard Stools/Constipation= High |
| Watt et al., 2024 (79) | Global  (23 studies) | Non anemic Pregnant women | oral iron supplementation | placebo or no supplement | **Birth outcomes**  (i)Caesarean section rates: No difference OR = 1.07; (0.88,1.29); p = 0.54;n=6  (ii)preterm births: No difference; OR = 0.82; (0.58,1.17); p = 0.28; n=6  (iii)Birth Weight: No difference ,MD 17.75g, (-55.74,91.24), p = 0.64  **GI side effects:**  No significant difference in nausea, vomiting, constipation, diarrhoea, abdominal pain, loss of appetite and heartburn. | ROB & NOS  “Low” only 2 studies, “Some concerns” for 13 studies and “High risk” for 6 studies |
| Hansen et al., 2023 (80) | Global  (08 studies) | Pregnant women | Daily oral iron supplements | placebo/ non iron supplementation | **Birth outcomes**  (i)LBW: Reduced incidence RR = 0.30; (0.13,0.68); n=2, 361  (ii) SGA: RR = 0.39; (0.17,0.86); n=1, 213).  (iii) preterm births: No difference RR = 0.90; (0.47,1.71); n=2, 361 | GRADE  Maternal ID at term, SGA, PTM= Low  Maternal IDA at term, LBW= Moderate  Maternal high Hbconcentrations duringpregnancy= Very Low |
| Peña-Rosas JP et al., 2015 (77) | Global  21 studies | Pregnant Women | daily iron and iron plus folic acid supplementation | Folic Acid/Placebo/ Vitamin Supplementation/Mineral Supplementation | No significant between iron supplementation and placebo/no iron groups: 25.3% vs. 9.91%.;Average risk ratio (RR) for side effects: 1.29; (0.83,2.02). n=11,2423 | GRADE |
| Petry et al., 2016 (71) | Global  90 studies | women (pregnant and lactating)  children (6–23 months) | Daily iron and zinc dose did not exceed 15 mg and 10 mg, respectively for children45 mg and 21 mg, respectively for women | Placebo/Non Iron Supplements | No beneficial impact of iron use on diarrhoea, respiratory infection and/or fever (8 studies, 11 comparison group) effect size(Meta analysis not done) | GRADE |

### Figure 1: PRISMA diagram of included studies
